# Characterization of direct and/or indirect genetic associations for multiple traits in longitudinal studies of disease progression

**DOI:** 10.1101/2021.05.10.21256880

**Authors:** Myriam Brossard, Andrew D. Paterson, Osvaldo Espin-Garcia, Radu V. Craiu, Shelley B. Bull

## Abstract

When quantitative longitudinal traits are risk factors for disease progression and subject to random biological variation, joint model analysis of time-to-event and longitudinal traits can effectively identify direct and/or indirect genetic association of single nucleotide polymorphisms (SNPs) with time-to-event. We present a joint model that integrates: *i)* a multivariate linear mixed model describing trajectories of multiple longitudinal traits as a function of time, SNP effects, and subject-specific random effects, and *ii)* a frailty Cox survival model that depends on SNPs, longitudinal trajectory effects, and subject-specific frailty accounting for dependence among multiple time-to-event traits. Motivated by complex genetic architecture of type 1 diabetes complications (T1DC) observed in the Diabetes Control and Complications Trial (DCCT), we implement a two-stage approach to inference with bootstrap joint covariance estimation and develop a hypothesis testing procedure to classify direct and/or indirect SNP association with each time-to-event trait. By realistic simulation study, we show that joint modelling of two time-to-T1DC (retinopathy, nephropathy) and two longitudinal risk factors (HbA1c, systolic blood pressure) reduces estimation bias in genetic effects and improves classification accuracy of direct and/or indirect SNP associations, compared to methods that ignore within-subject risk factor variability and dependence among longitudinal and time-to-event traits. Through DCCT data analysis, we demonstrate feasibility for candidate SNP modelling, and quantify effects of sample size and Winner’s curse bias on classification for two SNPs identified as having indirect associations with time-to-T1DC traits. Overall, joint analysis of multiple longitudinal and multiple time-to-event traits provides insight into complex trait architecture.

## INTRODUCTION

Despite their known ability to improve inference in clinical and epidemiological studies, particularly in the presence of informative censoring/dropout or when longitudinal traits are measured with biological random variation (Hogan and Laird 1998; Ibrahim et al. 2010; Chen et al. 2011), joint models for longitudinal and time-to-event (TTE) outcomes have received limited attention in genetic association study design and analysis. Genome-wide association studies (GWAS) of quantitative traits (QTs) often require follow-up analyses to identify whether SNP associations detected with each of the QT(s), analyzed separately, also affect related disease outcomes through direct and/or indirect effects induced by those QTs (Fig. 1). Such QTs can include established intermediate risk factors for clinical outcomes, which may be measured with high within-subject variability (e.g. random biological variation). By accounting for random measurement error in intermediate QT risk factors and dependencies among traits, joint model analysis can improve accuracy and efficiency of effect estimation, as well as detection and interpretation of SNP associations.

**Fig. 1.**
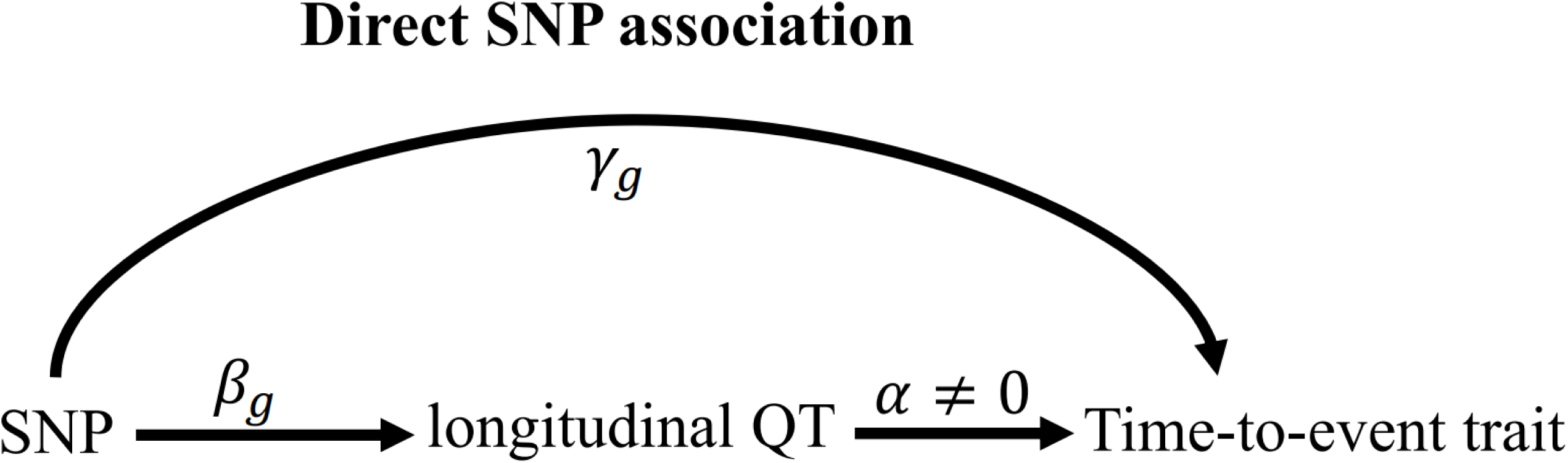
Directed acyclic graph (DAG) illustrating the joint model parameters to characterize the direct SNP effect on the time-to-event trait and the indirect SNP effect via the intermediate longitudinal QT associated with the time-to-event trait. Figure adapted from (Ibrahim et al. 2010) which proposed a general joint model formulation for one longitudinal QT and one time-to-event trait to address questions specific to testing for treatment effects in randomized-controlled clinical trials.

The multiple-trait extensions we develop stem from a random-effects joint model that consists of a sub-model for a single longitudinal trait linked to a sub-model for a single right-censored time-to-event trait (Wu et al. 2012; Asar et al. 2015). The longitudinal sub-model describes the QT as an underlying smooth trajectory that depends on fixed effects of time and baseline covariates, as well as subject-specific random effects. The joint model association structure is induced via the functional dependence between the hazard of an event at time *t* and the longitudinal trait trajectory (Hickey et al. 2016; Papageorgiou et al. 2019). Specification of this relationship can be based on prior biological knowledge of the link between the longitudinal and time-to-event traits. As previously elucidated, this class of joint models provides interpretations of direct and/or indirect effects because the relationship between a baseline covariate, such as a SNP genotype, and each of the longitudinal and time-to-event traits, as well as the relationship between the longitudinal and time-to-event traits can be specified via model parameters corresponding to direct, indirect and overall effects (Ibrahim et al. 2010). In this report, we demonstrate that extensions to jointly model multiple longitudinal and multiple time-to-event traits can further improve inference by *(i)* borrowing information shared among correlated traits, and *(ii)* accounting for indirect genetic pathways from multiple longitudinal QT risk factors, that if ignored, can confound an indirect genetic association with a direct association.

Joint model extensions have been reviewed for multiple longitudinal traits (Hickey et al. 2016; Papageorgiou et al. 2019) and for multiple time-to-event traits (Hickey et al. 2018a). Although a few extensions have been developed for both multiple longitudinal and multiple time-to-event traits, for example (Zhu et al. 2012; Tang et al. 2014; Tang and Tang 2014), these models are often formulated for a specific study question, and thus can lack generalizability. In addition, such extensions raise computational challenges for maximisation of the marginal likelihood that integrates over the distribution of the multivariate random effects. Two-stage approaches for joint model fitting are computationally efficient and allow more flexible model formulations (Self and Pawitan 1992; Tsiatis et al. 1995; Bycott and Taylor 1998; Dafni and Tsiatis 1998). However, in some circumstances, inference can be mis-calibrated when parameter estimates and predictions from Stage 1 are obtained from the longitudinal model without consideration of the time-to-event outcome, or when the uncertainty in Stage 1 estimates is ignored during Stage 2 estimation (Wulfsohn and Tsiatis 1997), a problem known as propagation of errors.

Motivated by the complex genetic architecture of long-term type 1 diabetes complications (T1DC), we develop a joint-model extension to evaluate genetic associations with multiple longitudinal QT risk factors and multiple TTE traits. Risk of development of T1DC, including diabetic retinopathy (DR) and diabetic nephropathy (DN), is hypothesized to result from multiple genetic factors with potential direct and/or indirect effects induced via multiple shared and/or specific QT risk factors (Paterson and Bull 2012). Besides potential genetic factors, hyperglycemia (measured by Hemoglobin A1c, hereafter abbreviated as HbA1c) represents a major risk factor for T1DC; intensive insulin therapy to control the HbA1c level to a normal range prevents and delays progression of long-term T1DC, as demonstrated by the Diabetes Control and Complications Trial (DCCT, (The Diabetes Control and Complications Trial Research Group 1993)). The first GWAS to analyze the DCCT study phenotypes identified two SNPs associated with within-patient mean HbA1c at genome-wide significance in the Conventional treatment arm, namely rs10810632 (in *BNC2*, 9p22.2) and rs1358030 (near *SORCS1,* 10q25.1), and reported weaker SNP associations with the secondary outcomes time-to-DR and/or time-to-DN (Paterson et al. 2010). Genetic association studies also reported variants with potential pleiotropic effects on DR and DN (Hosseini et al. 2015). Other measured longitudinal QTs, also influenced by genetic factors, are postulated to have associations with T1DC, for example, association of systolic blood pressure (SBP) with DN.

Our objective is to develop an integrated approach to investigate the complex genetic architecture of disease complications, and associated risk factors, as in the motivating study of type 1 diabetes complications. The approach we develop entails multiple longitudinal risk factors and multiple time-to-event outcomes, as well as multiple SNP associations with multiple traits. In addition to genetic variants associated with risk factors, the model needs to handle multiple longitudinal quantitative risk factors that can be related to more than one complication, and genetic variants that can affect risk of more than one complication directly and/or indirectly through intermediate risk factor(s); accounting for known intermediate longitudinal QTs is essential to correctly distinguish between direct and indirect genetic effects on each T1DC trait. To this end, we formulate a general model extension to multiple longitudinal QTs and multiple TTE traits, which proposes correlated random effects and a frailty term to address dependency among QTs and among TTE traits.

Based on the DCCT study, we hypothesize a multi-trait model for T1DC genetic architecture, and develop methods to investigate it. Because the goal of intensive therapy in DCCT was to reduce HbA1c into the non-diabetic range, which produced treatment differences in HbA1c values, we base our joint model evaluation and application on *N*=667 unrelated individuals of European ancestry from the Conventional treatment group. Longitudinal measurements for HbA1c and SBP were recorded irrespective of the occurrence of any complication event(s) at up to 39 quarterly visits (See File S1 for a description of the DCCT dataset we analysed). HbA1c and SBP are established risk factors for T1DC, and genetic association with either risk factor can induce an indirect genetic association with T1DC. The latter can be mistaken as a direct genetic association when the intermediate longitudinal risk factor(s) is ignored. Distinguishing between direct and/or indirect genetic effects can help to reveal genetic pathways in the aetiology of T1DC with implications for the direction of on-going investigations, and development of new intervention strategies. Nevertheless, accurate classification of direct and/or indirect SNP associations is challenged by within-patient variability in intermediate QT(s), and unmeasured shared risk factors among longitudinal and time-to-event traits.

The *primary* contribution of our work is a general formulation of a joint model for multiple longitudinal QT risk factors and multiple time-to-event traits in genetic association studies. We develop inference methods for statistical genetic analysis using a two-stage approach to joint model parameter estimation and hypothesis testing, including a procedure to classify SNP associations with each time-to-event trait as direct and/or indirect. A *second* contribution of this paper is the development of a data-informed simulation algorithm, under the postulated multi-trait model for T1DC genetic architecture, to generate multiple causal SNPs with various direct effects on simulated TTE traits and/or indirect effects via observed (measured) longitudinal QTs in DCCT and unobserved (simulated) longitudinal QTs. This algorithm provides a general approach to estimate power of the joint modeling approach (in comparison to alternative methods) given study sample size and various direct/indirect genetic associations via observed or unobserved longitudinal QT(s) risk factors for time to disease complications. Our numerical investigations show that the proposed method reduces estimation bias and improves accuracy of classification of direct and/or indirect SNP associations in comparison with separate joint models for each pair of longitudinal QT and time-to-event trait, and approaches that ignore measurement error in longitudinal QT(s). *Lastly*, we show computational feasibility and interpretation in an extended joint model application to DCCT genetic association analyses of candidate SNPs. Using the proposed procedure, we classify rs10810632 and rs1358030 as having indirect association with two T1DC traits via the HbA1c longitudinal risk factor, and obtain similar conclusions using alternative time-dependent association structures that account for cumulative and time-weighted effects of HbA1c on T1DC traits (Lind et al. 1995; Lind et al. 2010). Example programs written in R for data simulation and for application of the proposed joint model are available on GitHub.

## MATERIALS AND METHODS

### Model Formulation

We assume that a set of *M* SNPs have been genotyped, together with observation of *K* (1 *≤ k ≤ K*) unordered and non-competing time-to-event traits, such as multiple disease complications, and *L* (1 *≤ l ≤ L)* longitudinal QTs (*i.e.* intermediate risk factors) measured in *N* unrelated individuals indexed by *i* (1 ≤ *i* ≤ *N*). To characterize the genetic architecture of multiple longitudinal risk factors and multiple time-to-events, we formulate a shared-random-effects joint model that connects longitudinal and time-to-event sub-models through specified time-dependent association structures. For ease of presentation, we simplify the model notation by assuming no adjusting covariates but note that trait-specific and/or shared covariates, such as confounding factors or ancestry-related principal components can be easily incorporated. We first introduce the joint model for one longitudinal trait (*L* = 1) and one time-to-event trait (*K* = 1). Then, in the subsequent subsection, we present the extension for an arbitrary number of longitudinal and time-to-event traits.

#### Joint model for *one* longitudinal and *one* time-to-event trait

For each individual *i*, we define ***y_i_*** = (*y*_*i*,1_, …, *y*_*i*,*j*_, …, *y*_*i*,*J*_), as the vector of QT measures collected over the *J* visit times ***t_i_*** = (*t*_*i*,1_, …, *t*_*i*,*j*_, …, *t*_*i*,*J*_)*^T^* with 1 ≤ *j* ≤ *J* and *t*_*i*,1_ ≤ ⋯ ≤ *t*_*i*,*j*_ ≤ ⋯ ≤ *t*_*i*,*J*_. We denote (*T*_*i*_, *δ*_*i*_) as the vector of right-censored event time *T*_*i*_ and event indicator *δ*_*i*_ for the time-to-event trait, and assume 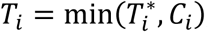, where 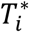 is the latent (uncensored) event time and *C*_*i*_ is the censoring time (e.g., administrative censoring). We define 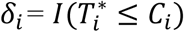, with *δ*_*i*_ = 1 if the event occurs during the observation period, and *δ*_*i*_ = 0 otherwise.

#### Longitudinal sub-model

This is specified by a mixed-effects model for the longitudinal QT, based on the (Laird and Ware 1982) linear mixed model. The model builds on the assumption that for every individual in the sample there exists an underlying smooth trajectory of the longitudinal QT that describes the subject-specific evolution dependent on time, SNP effect, and individual-level random effects ***b_i_***. To simplify the presentation, we assume a linear QT trajectory (Equation 1), but the longitudinal sub-model can be adapted for nonlinear trajectories using, for example, higher order polynomials or splines (Rizopoulos 2012):

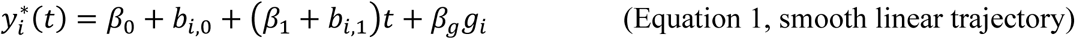

Where:

- *g*_*i*_, is the genotype of the individual *i* for the SNP being tested, coded as the number of copies of the minor allele,
- ***β*** = (*β*_0_, *β*_1_, *β_g_*)*^T^* is the vector of fixed intercept and slope time effects and fixed genetic effect on the longitudinal QT,
- ***b_i_*** = (*b*_*i*,0_, *b*_*i*,1_)*^T^* are the subject-specific random intercept and slope time effects assuming ***b_i_***∼*N*_2_(0, ***D***) and ***D*** is the variance-covariance matrix.

This trajectory cannot be observed directly, rather we observe longitudinal measurements ***y_i_*** collected at discrete time points *t*_*i*_ ; measurements are subject to independent and identically distributed noise contamination variables ***ε_i_***∼*N*_*J*_(0, ***Σ***), where ***ε_i_*** = (*ε*_*i*,1_, …, *ε*_*i*,*j*_, …, *ε*_*i*,*J*_)^*T*^, ***Σ*** = *σ*^2^***I***_***J***_, with *σ*^2^, the residual variance of the QT:

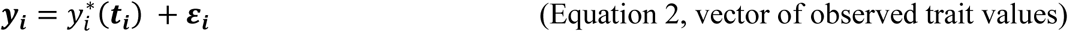

We assume that ***b_i_*** and ***ε_i_*** are independent (Laird and Ware 1982).

Equation 2 implies that ***y_i_*** in ℜ^*J*^ follows a multivariate normal distribution with:

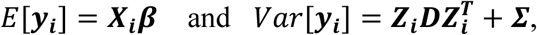

where ***X_i_*** = (**1**_***J***_, *t*_*i*_, *g*_*i*_**1**_***J***_) denotes the (*J-* by-3) design matrix for the fixed intercept, slope, and SNP effects, and ***Z***_*i*_ = (**1**_***J***_, *t*_*i*_) is the (*J-* by-2) design matrix for the random intercept and slope effects, with **1**_***J***_ = (1, …,1, …,1)^*T*^. To increase robustness to misspecification of the variance-covariance matrix ***D***, we adopt an unstructured form for the random-effects variance, defined as 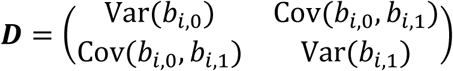, with the added benefit of not requiring additional constraints on the serial dependence between the repeated measurements for each individual. This choice implies that the covariance function between any pair of QT observations for individual *i* collected at two distinct visit times *t*_*i*,*j*_ ≠ *t*_*i*,*s*_ (1 ≤ *j* ≤ *J* and 1 ≤ *s* ≤ *J*, with *j* ≠ *s*) is given by *Cov*(*y*_*i*,*j*_, *y*_*i*,*s*_) = *t*_*i*,*j*_*t*_*i*,*s*_*Var*(*b*_*i*,1_) + (*t*_*i*,*j*_ + *t*_*i*,*s*_)*Cov*(*b*_*i*,0_, *b*_*i*,1_) + *Var*(*b*_*i*,0_) + *σ*^2^, with variance function 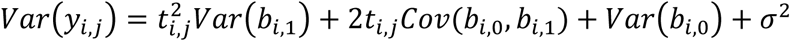, which is quadratic over time with positive curvature at *Var*(*b*_*i*,1_).

#### Time-to-event sub-model

This is specified by a proportional hazards model (PH model), in which the hazard function of the time-to-event trait is defined as the instantaneous event rate in a small interval around 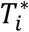 given that the event has not occurred before time *t*, genetic effect and a function of the history of the true unobserved longitudinal process up to time *t* that is associated with risk of the event, 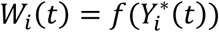, with 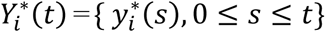. We specify that the hazard function (Equation 3) depends on the SNP effect adjusted for association of the longitudinal QT risk factor with the time-to-event trait.

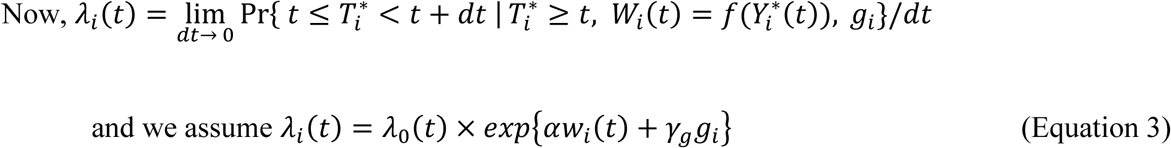

Where:

- *λ*_0_(*t*) is a (parametric or non-parametric) baseline hazard function;
- 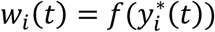 specifies the function of the longitudinal QT trajectory accounting for trajectory values at time *t* that is associated with risk of the event. In the case of a *contemporaneous* parametrization, the hazard of an event at a time *t* depends on the longitudinal trajectory value at the same time 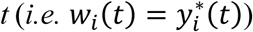. Other functional forms of the QT trajectory can weight earlier QT values according to prior knowledge of the relationship of the QT with the time-to-event trait (Hickey et al. 2016; Mauff et al. 2017; Papageorgiou et al. 2019);
- *α* is the effect of the longitudinal QT risk factor on the time-to-event trait;
- *γ*_*g*_ denotes the genetic effect on the time-to-event trait accounting for association of the longitudinal QT risk factor with the time-to-event trait.

#### Interpretation

As depicted in Fig. 1, the joint model parameters characterize relationships among a SNP, an intermediate QT, and a time-to-event trait and decompose possible effects of a SNP on a time-to-event trait into: an indirect effect induced via the SNP effect on the longitudinal QT; and a direct SNP effect independent of the QT (Ibrahim et al. 2010; Hickey et al. 2018b).

Based on the SNP effects, *β*_*g*_ and *γ*_*g*_, and assuming *α* ≠ 0, a SNP association with a time-to-event trait can be one of three types:

- indirect SNP association: the SNP has a non-null effect on the longitudinal QT (*β*_*g*_ ≠ 0), but no effect on the time-to-event trait (*γ*_*g*_ = 0); the overall SNP effect *θ* depends on the indirect effect (*θ* = µ_*g*_, with µ_*g*_ = *αβ*_*g*_).
- direct SNP association: the SNP has a non-null effect on the time-to-event trait (*γ*_*g*_ ≠ 0), but no effect on the longitudinal QT (*β*_*g*_ = 0); the overall SNP effect depends only on the direct effect (*θ* = *γ*_*g*_).
- both direct and indirect SNP associations: the SNP has non-null effects on the longitudinal risk factor (*β*_*g*_ ≠ 0) and on the time-to-event trait (*γ*_*g*_ ≠ 0). In this case, the overall SNP effect *θ* aggregates the indirect and direct SNP effects (*θ* = µ_*g*_ + *γ*_*g*_, with µ_*g*_ = *αβ*_*g*_).

In later subsections, we detail statistical implementation for estimation of the *α*, *β*_*g*_ and *γ*_*g*_ parameters and associated hypothesis testing in the joint model, which underlie the procedure we then propose to classify the SNP association as indirect, direct, or both direct and indirect.

When an associated longitudinal risk factor is omitted from the time-to-event model, the estimated SNP effect on the time-to-event trait captures the overall SNP effect (*θ* = µ_*g*_ + *γ*_*g*_). This can occur in GWAS when the time-to-event analysis ignores an intermediate risk factor or when the time-to-event trait is associated with more than one intermediate risk factor. This observation also illustrates one of the limitations of the joint model for *one* longitudinal with *one* time-to-event trait, with the consequence that an indirect SNP association can be mistaken as a direct association when other longitudinal risk factors are omitted in the joint model.

#### Generalization of the joint model to *multiple* longitudinal and *multiple* time-to-event traits

To characterize the genetic architecture of a system of multiple longitudinal risk factors and multiple time-to-events, we propose an extension of the joint model to *L* longitudinal and *K* time-to-event traits (*L>*1, *K>*1), as shown in Fig. 2 and detailed as follows. We define ***y***_*i*,***l***_ = (*y*_*i*,*l*,1_, …, *y*_*i*,*l*,*j*_, …, *y*_*i*,*l*,*J*_) as the observed longitudinal measures for each *l*^th^ QT, 1 ≤ *l ≤ L*, collected over the *J* visit times ***t_i_***. We define (*T*_*i*,*k*_, *δ*_*i*,*k*_) as the vector of observed right-censored event time *T*_*i*,*k*_ and event indicator *δ*_*i*,*k*_ for each *k^th^* time-to-event trait for individual *i*, with 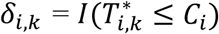. We assume the same censoring time *C*_*i*_ across all *K* outcomes, but the model can be extended to situations where *C*_*i*_ varies for each time-to-event trait. Again, for ease of presentation, we simplify the model notation with no adjusting covariates and assume linear trajectories for all *L* longitudinal traits and contemporaneous effects of *L* longitudinal traits on the *K* time-to-event traits. The model can be extended to account for non-linear trajectories, cumulative longitudinal effects, and trait-specific and/or shared covariates such as confounding factors or ancestry-related principal components.

**Fig. 2.**
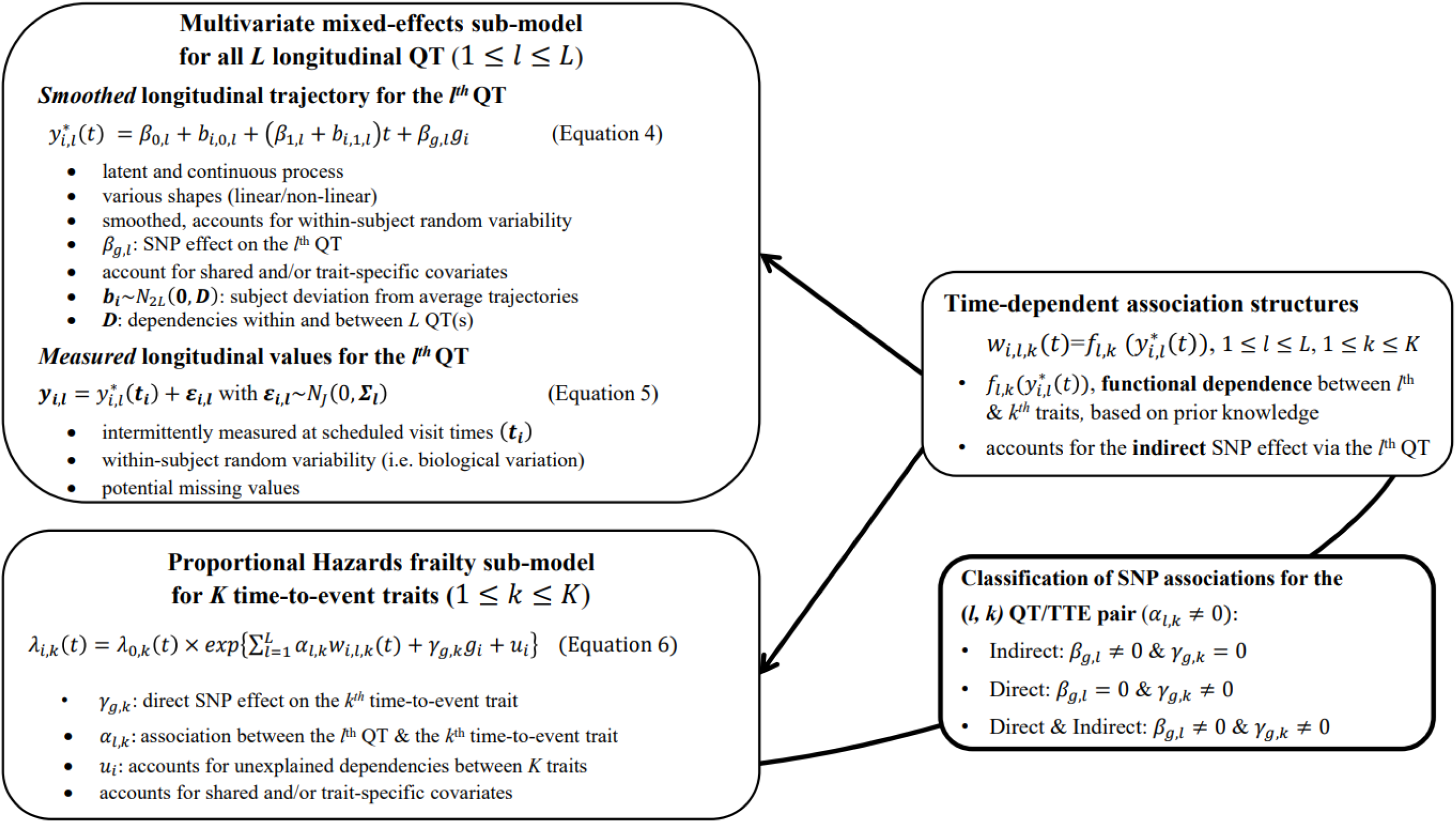
Proposed joint modelling approach for characterization of complex genetic architecture of multiple disease progression.

#### Multivariate longitudinal sub-model

In the extension of the longitudinal sub-model to *L* longitudinal traits, we index subscripts in Equations 1 and 2 for each *l*^th^ longitudinal trait (Equations 4 and 5 in Fig 2). The vector of observed trait values becomes:

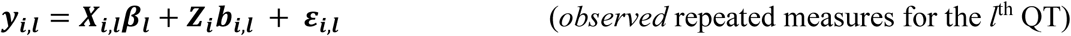

Where:

- ***X***_*i*,***l***_ = (**1**_***J***_, *t*_*i*_, *g*_*i*_**1**_***J***_) and ***Z***_*i*,***l***_ = (**1**_***J***_, *t*_*i*_) are the design matrices for fixed and random effects,
- ***β***_***l***_ = (*β*_0,*l*_, *β*_1,*l*_, *β*_*g*,*l*_)*^T^* and ***b***_*i*,***l***_ = (*b*_*i*,0,*l*_, *b*_*i*,1,*l*_)^*T*^ denote the QT-specific fixed and random effects,
- ***ε***_*i*,***l***_ = (***ε***_*i*,**1**,***l***_, …, ***ε***_*i*,***J***,***l***_, …, ***ε***_*i*,***J***,***l***_)^***T***^ is the vector of residual error terms, with ***ε***_*i*,***l***_∼***N***_***J***_(**0**, ***Σ***_***l***_), where 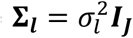 with 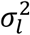, the residual variance for the *l*^th^ QT; we assume the ***ε***_*i*,***l***_ are independent for all *L* traits.

To account for dependence among the *L* longitudinal QTs, we assume the overall random effects vector for all *L* QTs, ***b_i_*** = (***b***_*i*,**1**_, …, ***b***_*i*,***l***_, …, ***b***_*i*,***L***_)^***T***^∼*N*_2*L*_(**0**, ***D***), where 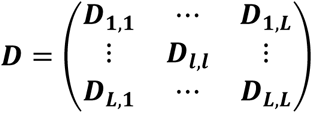 is the variance-covariance matrix for all *L* QTs, accounting for serial dependencies within each *l^th^* QT, i.e. 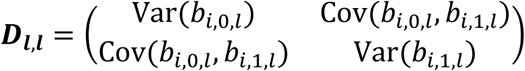, and accounting for cross-dependencies between each pair *l*, *m* of QTs with *l*≠*m,* that is 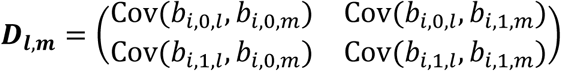.

This formulation implies that the vector of stacked repeated measures for all *L* longitudinal QTs for individual *i*, ***y_i_*** = (***y***_*i*,**1**_, …, ***y***_*i*,***l***_, …, ***y***_*i*,***L***_)^*T*^ in ℜ^*J*×*L*^ follows a multivariate normal distribution with mean *E*[***y_i_***] = ***X_i_β*** and variance 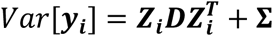, where:

- ***X_i_*** = *diag*(***X***_*i*,**1**_,…, ***X***_*i*,***l***_, …, ***X***_*i*,***L***_) and ***Z***_*i*_ = *diag*(***Z***_*i*,**1**_,…, ***Z***_*i*,***l***_, …, ***Z***_*i*,***L***_) are the overall (*JL*-by-3*L*) and (*JL*-by-2*L*) design block diagonal matrices for the fixed and random effects respectively;
- ***β*** = (***β***_**1**_, …, ***β***_***l***_, …, ***β***_***L***_)^*T*^ is the 3*L*-length stacked vector of fixed effects,
- ***D*** is the (2*L-* by*-*2*L*) covariance matrix for random effects ***b*** = (***b***_*i*,**1**_, …, ***b***_*i*,***l***_, …, ***b***_*i*,***L***_)^***T***^,
- **Σ** = *diag*(***Σ***_**1**_, …, ***Σ***_***l***_, …, ***Σ***_***L***_)^*T*^ is the (*JL*-by-*JL*) block diagonal matrix of residual variances.

Assuming, for each *l^th^* QT, an unstructured variance-covariance matrix ***D***_***l***,***l***_, the variance at each visit time *t*_*i*,*j*_ is *Var*(*y*_*i*,*j*,*l*_) and the covariance function *Cov*(*y*_*i*,*j*,*l*_, *y*_*i*,*s*,*l*_) between two visit times *t*_*i*,*j*_ ≠ *t*_*i*,*s*_ are analogous to those defined above for the joint model with a single longitudinal and single time-to-event trait. The multivariate mixed model accounts for dependencies between each QT pair *l* ≠ *m* via random-effect covariance functions in the ***D***_***l***,***m***_ matrices where the covariance between observations of two QTs (*l*, *m*; *l* ≠ m) measured at the same visit time *t*_*i*,*j*_ is 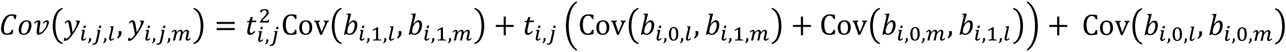 which is quadratic over time, and the covariance function between two longitudinal QTs (*l* ≠ *m*) measured at different visit times 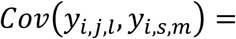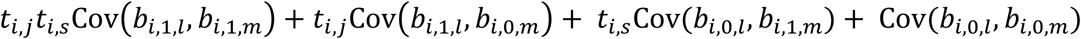. Thus, joint analysis of correlated longitudinal QTs is expected to improve power over separate analysis of each QT by borrowing information through implied dependency structures among the random effects.

#### Multivariate time-to-event sub-model

Finally, we extend Equation 3 to a multivariate PH frailty time-to-event sub-model, with a subject-specific random effect (frailty term, *u*_*i*_) introduced to capture potential unexplained dependencies (e.g. due to unmeasured baseline shared factors) among the time-to-event traits. In Equation 6 (Fig 2), *λ*_0,*k*_(*t*) and *γ*_*g*,*k*_, correspond to the baseline hazard function, and SNP effect on the *k*^th^ time-to-event trait (1 *≤ k ≤ K*), accounting for association of each *l*^th^ QT with the time-to-event trait *k* (*α*_*l*,*k*_, 1 ≤ *l* ≤ *L*). We assume that the subject-specific frailty term follows a gamma distribution, that is *u*_*i*_∼Γ(*a*, *b*) with *a*, *b*>0, and represents dependencies among the *K* traits. Equation 6 (Fig 2) can be expressed as:

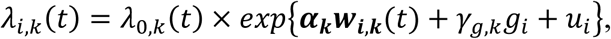

where ***α***_***k***_ = (*α*_1,*k*_, …, *α*_*l*,*k*_, …., *α*_*L*,*k*_)^*T*^ is the vector of all *L* QT effects on the *k*^th^ time-to-event trait, and ***w***_*i*,***k***_(*t*) = (*w*_*i*,1,*k*_(*t*), …., *w*_*i*,*l*,*k*_(*t*), …, *w*_*i*,*L*,*k*_(*t*)) specifies the corresponding association profile of each QT with the *k*^th^ time-to-event trait. We note 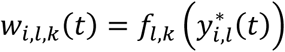, where 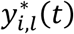, denotes the *l*^th^ QT trajectory (1 ≤ *l* ≤ *L*) at time *t*, which depends on the fixed and random effects ***β***_***l***_ and ***b***_*i*,***l***_.

#### Comparisons with joint model of one longitudinal and one time-to-event trait

In the proposed joint model extension for multiple longitudinal and multiple time-to-event traits, the direct, indirect and overall SNP effects defined above for the joint model with one longitudinal and one time-to-event trait are interpreted similarly. However, there are important practical differences between the latter model for a pair of traits and the proposed multi-trait. *First*, because the joint model extension can account for multiple intermediate longitudinal QT risk factors associated with one (or multiple) time-to-event trait(s), it improves inference for SNP association and accuracy of SNP classification, particularly when a time-to-event trait depends on more than one longitudinal risk factor as illustrated in our numerical experiments. *Second,* in the multivariate longitudinal sub-model, the variance-covariance matrix ***D*** for the random effects specifies non-null covariance terms in ***D***_***l***,***m***_ for each pair of longitudinal QT *l*, *m* (1 ≤ *l* ≤ *L* and 1 ≤ *m* ≤ *L*, *l* ≠ *m*). In contrast, under the assumption of null covariance terms in ***D***_***l***,***m***_ for all QT pairs inherent in separate analyses of each QT, the multivariate sub-model reduces to independent sub-models for each longitudinal QT. When longitudinal QTs are correlated, assuming null covariances can fail to make use of information borrowed through the random effects and reduce efficiency of the parameter estimates in the longitudinal trajectories (Shah et al. 1997; Jensen and Ritz 2018). This, in turn, can affect estimation in the time-to-event model. *Third*, without a frailty term *u*_*i*_, the time-to-event sub-model (Equations 6 and 7) reduces to separate time-to-event sub-models for each time-to-event trait. *Thus*, through use of a shared frailty term, the extended joint model accounts for residual dependency between the *K* time-to-event traits, not explained by the covariates shared by the time-to-event sub-models. *Overall*, the proposed joint model for multiple longitudinal and multiple time-to-event traits can improve inference by accounting for intermediate longitudinal QT(s) and their dependencies, as well as dependencies among the time-to-event traits, and thereby improve classification accuracy of direct and/or indirect SNP associations.

### Implementation

#### Effect estimation and test statistic construction

To address computational obstacles involved in the maximization of the joint likelihood and allow more flexible inference, we estimate the parameters using a two-stage approach (see details in the Appendix). We work within the framework originally defined by (Tsiatis et al. 1995; Wulfsohn and Tsiatis 1997; Dafni and Tsiatis 1998; Tsiatis and Davidian 2001) and in the spirit of subsequent authors (Ye et al. 2008; Yuen et al. 2018; Arisido et al. 2019); (Tsiatis and Davidian 2001; Tsiatis and Davidian 2004) specify conditions that guarantee the two-stage estimators are consistent and asymptotically normal. Specifically, in Stage 1 we fit a multivariate mixed model (Equation 5) using the *mvlme*() function from the R package JoineRML [(Hickey et al. 2018c), version 0.4.2] to estimate the parameters of the longitudinal trajectories of the risk factors, and obtain fitted values of the smoothed trajectories. In Stage 2, we fit a Cox PH frailty time-to-event model (Equations 6 or 7) adjusting for functions of the smoothed trajectories as time-dependent covariates using the *coxph*() function from the R package survival [(Therneau and Grambsch 2000; Therneau 2020), versions 3.2.7 and 3.2.13]. We assume a Gamma distribution for the frailty term (*u*_*i*_) and a separate non-parametric baseline hazard function for each time-to-event trait using the strata argument in *coxph*().

To account for propagation of errors, due to uncertainty in Stage 1 estimates that is not accounted for during Stage 2 parameter estimation (Wulfsohn and Tsiatis 1997), and to empirically estimate the joint covariance matrix of SNP-QT trajectory (*β*_*g*,*l*_) and SNP-TTE effects (*γ*_*g*,*k*_), we apply a nonparametric bootstrap. The bootstrap also provides reliable standard error estimates in the joint time-to-event sub-model needed when using an unspecified baseline hazard (Hsieh et al. 2006; Lawrence Gould et al. 2015; Furgal et al. 2019). For each bootstrap sample *b* (1 ≤ *b* ≤ *B, B* is the total number of bootstrap repetitions), we generate a new dataset by randomly sampling *N* individuals with replacement and refitting the joint model on each new dataset *b*. We compute the empirical joint covariance matrix for all *β*_*g*,*l*_, *γ*_*g*,*k*_ and *α*_*l*,*k*_ parameter estimates using the *B* bootstrap parameter vector estimates. Wald statistics for each *β*_*g*,*l*_ are computed as 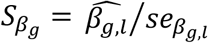 using the empirical bootstrap standard errors *se*_*βg*,*l*_ (to test H0: *β*_*g*,*l*_ = 0 *vs* H1: *β*_*g*,*l*_ ≠ 0), and similarly for each *γ*_*g*,*k*_ as 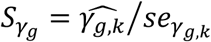 (to test H0: *γ*_*g*,*k*_ = 0 *vs* H1: *γ*_*g*,*k*_ ≠ 0).

In contrast to the two-stage joint model, a conventional one-stage analysis to assess whether a SNP has an association with the time-to-event trait, independent from the QT-TTE association, relies on regression adjustment using observed longitudinal QT values as time-dependent covariates in a Cox-PH model (Paterson et al. 2010; Deng and Pan 2017). This approach, based only on the time-to-event model, does not provide information about SNP-QT association (*β*_*g*,*l*_) and interprets the SNP as having a direct association with the time-to-event if the test of SNP-TTE effect (*γ*_*g*,*k*_) is declared significant, given the observed QT. Inference for *α*_*l*,*k*_ under this approach can be biased or inefficient when the QT is measured with random error or high within-subject variability (Faucett and Thomas 1996; Wulfsohn and Tsiatis 1997; Xu and Zeger 2001; Song et al. 2002; Brown and Ibrahim 2003), and inference for *γ*_*g*,*k*_ may also be affected. Although estimates of the SNP-QT effect (*β*_*g*,*l*_) obtained from mixed model QT analysis, fitted separately, may be used to distinguish between direct alone versus both direct and indirect SNP association, unlike the joint model, this two-step conditional approach ignores measurement error in the observed QT values.

#### Procedure to classify direct and/or indirect SNP associations

In Table 1, we present a practical procedure to classify a SNP as having direct and/or indirect association with a time-to-event trait *k,* that accounts for the SNP association with a longitudinal risk factor *l*. This procedure requires two significance thresholds, 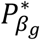 and 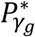 for hypothesis tests of each of *β_g,l_* and *γ_g,k_* respectively, to be specified prior to the analysis and adjusted for the number of SNPs tested. Depending on the research question, we can choose different values for 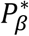 and 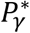, or the same value 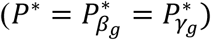. The latter is applicable, for instance, to systematically classify direct and/or indirect association for a set of *M* SNPs, and the former to assess which SNPs, among those reported to be associated with the longitudinal risk factor, have a direct effect on a time-to-event trait. To our knowledge, no comparable procedure to classify direct and/or indirect SNP association, based on SNP effect estimates from joint models, has been proposed for studies with longitudinal risk factors and time-to-event traits. A key feature of the proposed joint model extension to multiple longitudinal and multiple time-to-event traits is inference for SNP effects on each of the traits in a single integrated statistical model, while accounting for within-subject QT variability and dependencies among the traits. The focus of the simulation study and DCCT data application, which follow, is to evaluate the SNP classification procedure applied in extended joint model analysis.

**Table 1.** Procedure to classify a SNP as having an association with a time-to-event trait *k*, indirectly through an associated longitudinal risk factor *l* and/or directly with trait *k*, based on hypothesis tests of SNP effects *β_g,l_* and *³_g,k_*.

| | | SNP association with the longitudinal risk factor $l$ | |
| --- | --- | --- | --- |
| | | $P_{\beta_{g,l}} \leq P_{\beta_g}^*$ | $P_{\beta_{g,l}} > P_{\beta_g}^*$ |
| SNP association with the time-to-event trait $k$ | $P_{\gamma_{g,k}} \leq P_{\gamma_g}^*$ | Direct & Indirect<br>$\beta_{g,l} \neq 0$ AND $\gamma_{g,k} \neq 0$ | Direct<br>$\beta_{g,l} = 0$ AND $\gamma_{g,k} \neq 0$ |
| | $P_{\gamma_{g,k}} > P_{\gamma_g}^*$ | Indirect<br>$\beta_{g,l} \neq 0$ AND $\gamma_{g,k} = 0$ | Not Direct & Not Indirect<br>$\beta_{g,l} = 0$ AND $\gamma_{g,k} = 0$ |

$P_{\beta_{g,l}}$ and $P_{\gamma_{g,k}}$ are $P$ -values from Wald tests (1df) for each of SNP effects $\beta_{g,l}$ and $\gamma_{g,k}$ . For the former, we test $H_0: \beta_{g,l} = 0$ vs $H_1: \beta_{g,l} \neq 0$ . For the latter, $H_0: \gamma_{g,k} = 0$ vs $H_1: \gamma_{g,k} \neq 0$ . $P_{\beta_g}^*$ and $P_{\gamma_g}^*$ are the corresponding classification thresholds (see Materials and Methods for details). For example, if the $H_0: \beta_{g,l} = 0$ is rejected and $H_0: \gamma_{g,k} = 0$ is rejected by the corresponding test statistics, then the SNP is classified as being both indirectly and directly associated with the time-to-event trait $k$ . The classification procedure is based on the two separate test statistics and does not require a joint test statistic of the overall SNP effect; it thus partitions the two-dimensional parameter space into four mutually exclusive quadrants. In the simulations and application, we use $P^* = P_{\beta_g}^* = P_{\gamma_g}^*$ , although different thresholds can be specified for $P_{\beta_g}^*$ and $P_{\gamma_g}^*$ .

## Simulation study

### Design of the DCCT-data-based simulation study

To assess parameter estimation accuracy, hypothesis testing for tests of each genetic effects under the proposed joint model, and evaluate accuracy of the procedure we propose to classify SNPs as directly and/or indirectly associated with a time-to-event trait, we generate *R=*1000 replicated datasets simulated under a *complex genetic architecture* informed by the DCCT Genetics Study data (Fig. 3). The latter involves: *N*=667 subjects from the Conventional treatment group, *M*=5 simulated causal SNPs with direct effects on *K*=2 simulated time-to-T1DC (with ∼54% DR events and ∼25% DN events on average) and/or indirect effects via *L*=3 longitudinal QTs: two as measured in DCCT (HbA1c, SBP) and another simulated QT (*U*) that is unmeasured and designed to induce shared dependency among the T1DC traits. We assume effects of sex on SBP, and effects of T1D duration (at baseline) on both T1DC traits, as estimated in the original DCCT data, and specify contemporaneous association structures for the association of HbA1c and SBP on T1DC traits. We specify SNP effects and minor allele frequencies (MAFs) of genetic associations, as well as other parameter values according to the DCCT Genetics Study and the T1DC literature (Fig. 3). For SBP and DN, we inflate the typical SNP effect sizes observed in the literature to achieve power sufficient to detect SNP associations given the available DCCT sample size. Under the *global null genetic* scenario in which none of the SNPs is associated with any traits, we also simulate *M* SNPs with the same MAFs as for the causal SNPs, independently of the traits.

**Fig. 3.**
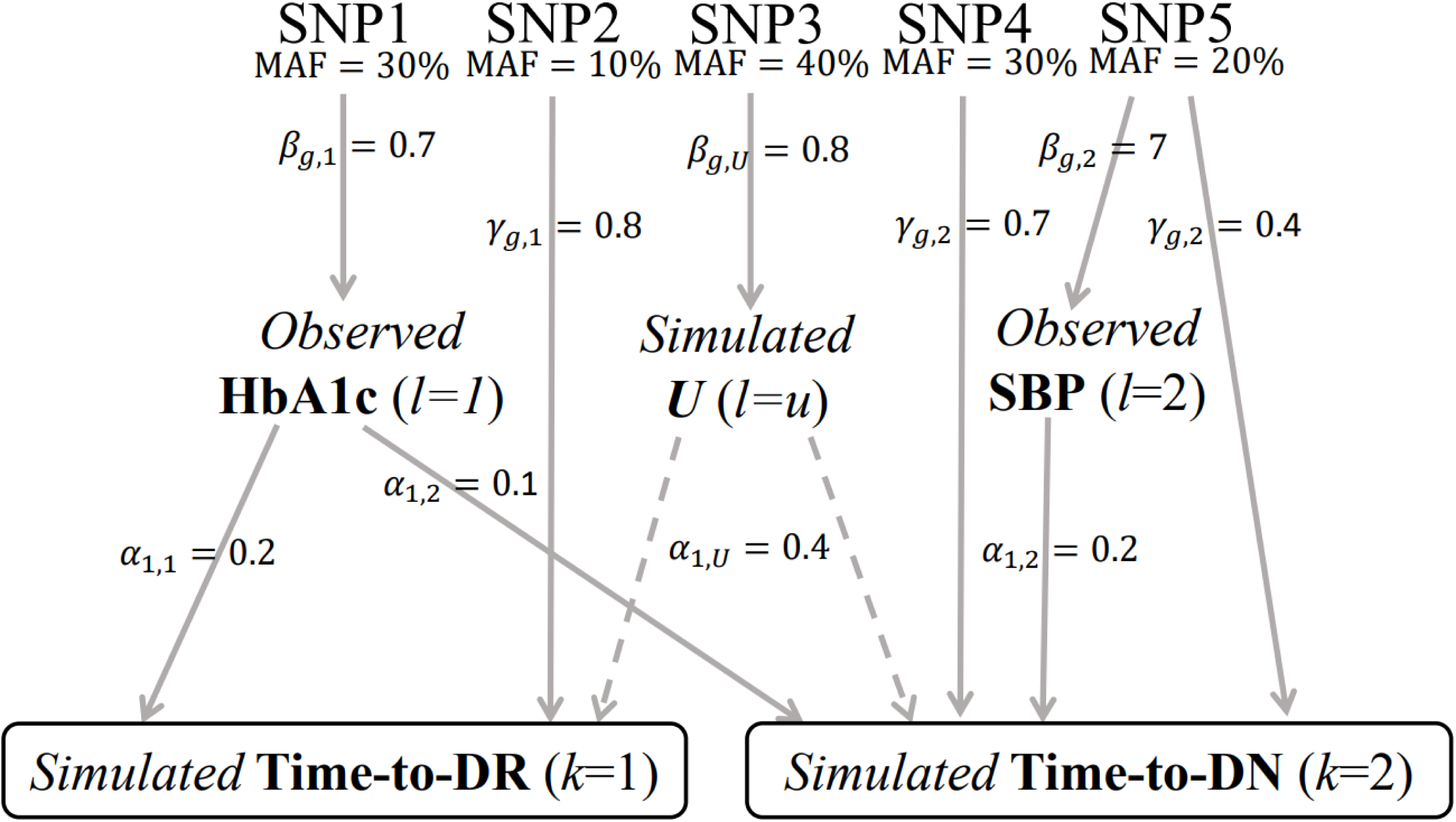
Realistic DCCT-data-based causal genetic scenario. We generated *R*=1000 replicates of *N*=667 DCCT individuals with *M*=5 causal variants and *K*=2 time-to-event traits simulated under this causal genetic scenario, and *R*=1000 replicates of *M*=5 SNPs (with same MAF as the causal ones) simulated under a *global null* genetic scenario where none of the SNPs is associated with any traits. The effects of gender on SBP, and of T1D duration at baseline on both time-to-T1DC traits are not represented in this figure, but are included in the data generating model, see File S2 (sections 2 and 3) for details.

#### Algorithm for realistic data generation under a complex genetic architecture

To generate a data structure that combines observed and simulated traits, we formulate a genotype-phenotype multi-trait model including: *(i) L=3* linear mixed models linking each SNP with an indirect effect to a longitudinal risk factor, and *(ii) K=2* non-independent parametric time-to-event models depending on fitted longitudinal QT trajectories and SNPs with direct effects. For each DCCT individual *i* with observed longitudinal measures for HbA1c and SBP, and observed baseline covariates (T1D duration, sex), we simulate: genotypes at *M* causal SNPs with MAF vector ***p***, longitudinal trait values ***U***_*i*_, and time-to-event traits ((*T*_*i*,*k*_, *δ*_*i*,*k*_), *k*=1,2 for DR and DN), using the algorithm illustrated in Fig. 4 and detailed in File S2 (sections 1-5). All SNP genotypes are generated under Hardy-Weinberg and linkage equilibrium assumptions. SNPs with indirect effects through longitudinal QTs associated with DR and/or DN are generated from the observed (SNP1, SNP5) or simulated (SNP3) QTs, while SNPs with direct effects (SNP2, SNP4) are generated independently of the longitudinal QTs and are included in the hazard function used to generate each time-to-event trait (Fig. 4).

**Fig. 4.**
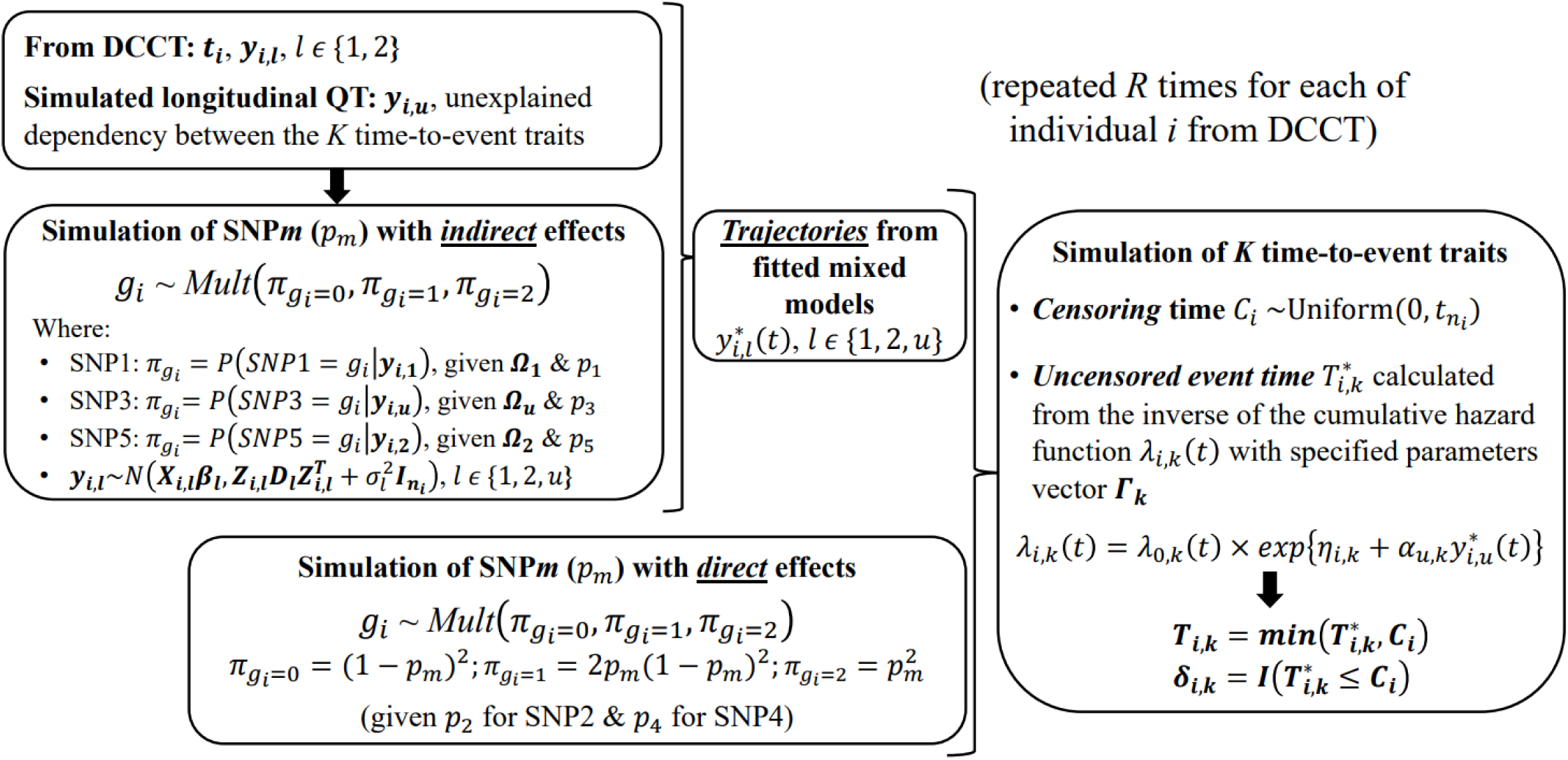
Illustration of the procedure developed for DCCT-based simulation study under the scenario from Fig. 3. For each individual *i,* with {*t*_*i*_, ***y***_*i*,**1**_, ***y***_*i*,**2**_} observed in DCCT, the algorithm simulate: latent longitudinal QT values for *U* (***y***_*i*,***u***_), genetic data at *M* causal SNPs (with ***p***, the specified vector of MAFs) and time-to-T1DC data (*T*_*i*,*k*_, *δ*_*i*,*k*_) for *K*=2 time-to-T1DC traits. The genetic data are simulated under Hardy-Weinberg and linkage equilibrium assumptions. SNPs with indirect effects are simulated from a multinomial distribution with calculated conditional genotype probabilities for individual *i* (***π***_*gi*=0_, ***π***_*gi*=1_, ***π***_*gi*=2_) based on ***y***_*i*,***l***_. Each ***y***_*i*,***l***_ is assumed to follow a multivariate normal distribution with ***X***_*i*,***l***_ and ***Z***_*i*,***l***_ the specified fixed and random effect design matrices in longitudinal trait models and 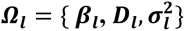 the vector of specified parameter values for each *l*^th^ QT. SNPs with direct effects are simulated from the population probabilities, that depend only on the MAF. The specified hazard function for each *k^th^* time-to-event trait depends on the effects of the longitudinal QT trajectories and on the SNPs with direct effects in *η*_*i*,*k*_, with 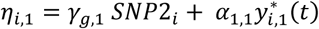 for DR and 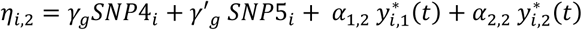 for DN, as well as the effect of the shared latent QT trajectory 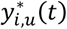 used to induce some dependencies between the time-to-event traits. We define ***Γ***_***k***_ as the vectors of specified parameter values for each *k*^th^ time-to-event trait. The uncensored event time 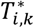, is simulated by calculating the inverse of the cumulative specified hazard function using the Brent univariate root-finding method (Brent 2013; Crowther and Lambert 2013). To simplify the exposition of the simulation procedure, we ignore the effects of the sex on SBP and of the T1D duration on both T1DC traits, but they were included in the data generating model, see File S2 for details. Parameters for the causal genetic scenario are shown in Fig. 3 and File S2 (section 4)

#### Scenario for DCCT-based complex genetic architecture

Overall, the complex genetic architecture represents multiple types of SNP-trait associations (Fig. 3): direct association with each T1DC trait (SNP2, SNP4), indirect association with both T1DC traits via measured (SNP1) and unmeasured (SNP3) longitudinal QTs; and direct and indirect association via a measured longitudinal QT (SNP5); all longitudinal risk factors exhibit within-subject random variability. Except for SNP3, all other SNP scenarios represent SNP association with a longitudinal QT risk factor (SNP1, SNP5) or a time-to-event trait (SNP2, SNP4, SNP5) testable in a single-trait GWAS. SNP1, SNP3 and SNP5 have indirect effects on T1DC traits, such that their associations with the T1DC traits are detectable in discovery analysis for each TTE trait, using Cox PH time-to-event models fitted separately for each TTE trait and ignoring the longitudinal QT risk factors (File S2, section 8). SNP1 corresponds roughly to rs10810632 and rs1358030 associations reported in the motivating DCCT GWAS of HbA1c (Paterson et al. 2010), while SNP5 represents a strong signal that would be detected in separate GWAS analysis of each longitudinal and time-to-event trait.

#### Analysis of the simulated data

To evaluate the statistical performance under the complex genetic architecture outlined above, focusing on direct and/or indirect classification, we compare extended joint model analysis of multiple longitudinal QT risk factors and multiple T1DC with alternative analysis methods that do not fully exploit the data structure. These alternative approaches include joint model analyses limited to two longitudinal QT and one time-to-event trait, joint model analyses of one longitudinal QT and one time-to-event trait, and a conditional time-to-event analysis of two time-to-event traits adjusted for observed values of two longitudinal QTs. We assess the impact of model specification on the classification accuracy of direct and/or indirect SNP associations by fitting mis-specified joint models that leave out important covariates or traits. Altogether, the comparisons are designed to assess the merits of extended joint model analysis over simpler available methods. In each replicated dataset, each of the five SNPs is separately analysed for association with the longitudinal QTs and TTE traits, using four alternative analytic approaches:

- JM-cmp: a *completely* specified joint model analysis that includes observed (HbA1c, SBP) and unobserved (*U*) longitudinal QT as well as baseline covariates (sex, T1D duration) used in the data simulation. Due to the latent nature of unobserved *U*, JM-cmp cannot be fitted in practice, but we include it as a benchmark for comparison against the data analysis models JM-mis that are fitted without *U*.
- JM-mis: includes the same variables as in JM-cmp, but excludes *U*, the unobserved longitudinal QT.
- JM-sep: joint models of *two* longitudinal traits and *one* time-to-event trait that do not account for dependency between the time-to-event traits (where JM-sep (*l* = 1,2; *k*=1) denotes the joint model for DR; and JM-sep(*l* = 1,2; *k*=2) the joint model for DN), and joint models of *one* longitudinal and *one* time-to-event trait that do not account for dependence between the longitudinal traits, nor between the time-to-event traits (referred to as JM-sep(*l* = 1; *k*=1), JM-sep(*l* = 1; *k*=2), and JM-sep(*l* = 2; *k*=2)). Altogether, the JM-sep models assess the merits of the extended joint model methods in comparison to JM-mis and JM-cmp.
- CM-obs: a Cox PH frailty survival analysis of both time-to-event traits (DR, DN) that includes the same variables as in JM-mis but adjusts for the *observed* longitudinal QT values as time-dependent covariates; this model corresponds to the conditional analysis approach mentioned above. Here, to classify SNPs as indirect association, we fit a linear mixed model for both QTs to test the SNP effects on the QT(s) adjusted for the same covariates as used for the joint models. Comparisons of estimation, hypothesis testing, and classification results based on CM-obs to those based on JM-mis and JM-cmp allow us to assess the impact of within-subject QT variation/measurement errors on hypothesis testing and classification results for each SNP.

For each of these analyses, we compute empirical covariance matrices for the effect estimates using 500 bootstrap iterations and construct large sample test statistics for each of the SNP effect parameters. Under two-stage JM inference, in stage 1 we fit a bivariate (*l*=1,2) or univariate (*l*=1 or *l*=2) longitudinal QT model and test the *β*_*g*,*l*_ parameters for indirect SNP association. Then in stage 2, we fit the time-to-event models with the QT trajectories from stage 1 according to the TTE model specification (*k*=1,2; *k*=1, or *k*=2), and test the *γ*_*g*,*k*_ parameters for direct SNP association. Because stage 1 QT analysis is shared among methods, differences in classification among analytic methods largely arise through differences in the test results in stage 2 TTE analysis.

Under two-stage and conditional independence assumptions described in an Appendix, the SNP association estimates of *β*_*g*,*l*_ and *γ*_*g*,*k*_ and corresponding test statistics are expected to be uncorrelated under the *global null* scenario, but this may not necessarily hold under the *genetic alternative* scenario when the analysis model is mis-specified or when the time-to-event estimation uses observed longitudinal trait values. We therefore compute empirical correlations in each replicate under both scenarios.

Given hypothesis test results for a pair of QT/TTE traits for each SNP, we apply the procedure defined in Table 1 to classify the SNP-TTE association.

#### Evaluation criteria

We compare type I error and power of hypothesis tests of SNP-QT (*β*_*g*,*l*_) and SNP-TTE (*γ*_*g*,*k*_) association among the alternative analytic approaches (JM-cmp, JM-mis, JM-sep, CM-obs) under the *global null* and *causal* genetic scenarios for each of the 5 SNPs analysed separately, at *P*\* = 5% and 1% critical values. We assess estimation accuracy using mean bias for *β*_*g*,*l*_ and *γ*_*g*,*k*_ estimates, and confidence interval coverage across replicates, and similarly examine the distribution and mean of their bootstrap standard errors and correlation for all the compared models.

For each of the 5 SNPs, we evaluate accuracy of the procedure presented in Table 1 to classify SNP association with each of the TTE traits as direct and/or indirect. Specifically, under the *global null* and *causal genetic* scenarios, we compare the empirical classification frequencies to the expected classification frequencies under the assumption of indirect and/or direct association built into the generating model. The empirical frequencies are tabulated from the distribution of simulation replicates in the four classification categories (direct, indirect, direct and indirect, not direct and not indirect) as defined in Table 1, using specified classification thresholds such that 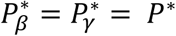, with *P*;^*^=0.05. We calculate expected frequencies under the assumption that the ***Z*** statistics, constructed from the estimates of *β*_*g*,*l*_ and *γ*_*g*,*k*_, and their bootstrap variances, follow a bivariate normal distribution with correlation specified by the bootstrap estimates (see File S2, section 9 for details), to allow for potential dependence between the SNP parameter tests. We judge the classification procedure for a SNP association with each QT-TTE trait pair to have high accuracy when the empirical frequencies are consistent with those expected, and we compare accuracy among the different models. We also assess empirical classification frequencies for SNPs with non-null effects across variation in stringency of significance thresholds up to *P*;^*^=10^−5^.

## Availability

DCCT data are available to authorized users at https://repository.niddk.nih.gov/studies/edic/ and https://www.ncbi.nlm.nih.gov/projects/gap/cgi-bin/study.cgi?study_id=phs000086.v3.p1 (IRB #07-0208-E). Example R codes for DCCT-data-based simulation and analysis of the simulated data are provided on GitHub (https://github.com/brossardMyriam/Joint-model-for-multiple-trait-genetics). Supplementary files are available online (https://figshare.com/s/2b9f6b3da5e1f03e8086). File S1 includes the description of the DCCT dataset as well as the list of the participants of the DCCT/EDIC Research Group; File S2 includes supplemental information for the DCCT-based simulation study; File S3 includes supplemental information for the Analysis of the DCCT Genetics Study data; File S4 includes the list of SNPs analyzed in DCCT; File S5 includes some notes on a multi-trait SNP association test for SNP effects estimated under the proposed joint model framework. Computations were run on the Niagara supercomputer (see https://docs.computecanada.ca/wiki/Niagara#Niagara_hardware_specifications and https://docs.scinet.utoronto.ca/index.php/Niagara_Quickstart for the hardware specifications and characteristics). Computational resources and time used to fit the joint model of two quantitative traits (HbA1c, SBP) and two time-to-event traits (DR, DN) in DCCT for one SNP are provided as part of the discussion section.

## RESULTS

### Simulation Study

#### SNP association test validity and power

Under the *global null* scenario of no genetic association with any of the traits, the type I error of each SNP test (*β*_*g*,*l*_ and *γ*_*g*,*k*_) is reasonably well controlled (Table 2, left-hand side), and the *P*-values *P*;_*βg*_ and *P*;_*γg*_ from the joint models show no departure from the expected large sample distributions (*χ*^2^ with 1 degree of freedom, see section 7 in File S2).

**Table 2.** Empirical type-I error and power (%) for SNP hypothesis tests of each of β_g,l_ and γ_g,k_ based on the complete joint model and compared models, assessed using R=1000 replicates of N=667 DCCT subjects, with SNPs simulated under global genetic null and genetic alternative simulation scenarios. Values are computed for each SNP and each genetic association parameter as the proportion of replicates that reject the null hypothesis at significance threshold P*=0.05. Results at other significance levels P*, and also for CM-obs, are shown in File S2, under the global null scenario at P* ≤ 0.01 (section 7), and under the alternative genetic scenario at *P*;^*^ ≤ 10^−5^ (section 8).

| SNPs<sup>1</sup> | Analysis Models | Global null at $P^*=5\%$ | | | | Genetic alternative at $P^*=5\%$ | | | |
| --- | --- | --- | --- | --- | --- | --- | --- | --- | --- |
|  |  | HbA1c | SBP | DR | DN | HbA1c | SBP | DR | DN |
| SNP1<br>(MAF=30%) | | $\beta_{g,1}=0$ | $\beta_{g,2}=0$ | $\gamma_{g,1}=0$ | $\gamma_{g,2}=0$ | $\beta_{g,1}=0.7$ | $\beta_{g,2}=0$ | $\gamma_{g,1}=0$ | $\gamma_{g,2}=0$ |
|  | JM-cmp | 5.2 | 5.8 | 4.4 | 4.8 | **100** | 5.1 | 4.1 | 5.4 |
|  | JM-mis | 5.2 | 5.8 | 3.6 | 4.9 | **100** | 5.1 | 4.2 | 4.6 |
| | JM-sep($l=1,2; k=1$) | 5.2 | 5.8 | 3.8 | . | **100** | 5.1 | 4.7 | . |
| | JM-sep($l=1,2; k=2$) | 5.2 | 5.8 | . | 5.2 | **100** | 5.1 | . | 4.6 |
| | JM-sep($l=1; k=1$) | 5.6 | . | 4.0 | . | **100** | . | 4.6 | . |
| | JM-sep($l=1; k=2$) | 5.6 | . | . | 4.6 | **100** | . | . | 5.7 |
| | JM-sep($l=2; k=2$) | . | 5.9 | . | 4.6 | . | 5.2 | . | 11.2 |
| SNP2<br>(MAF=10%) | | $\beta_{g,1}=0$ | $\beta_{g,2}=0$ | $\gamma_{g,1}=0$ | $\gamma_{g,2}=0$ | $\beta_{g,1}=0$ | $\beta_{g,2}=0$ | $\gamma_{g,1}=0.8$ | $\gamma_{g,2}=0$ |
|  | JM-cmp | 5.5 | 6.0 | 5.8 | 3.9 | 6.6 | 5.4 | **100** | 3.9 |
|  | JM-mis | 5.5 | 6.0 | 4.5 | 3.8 | 6.6 | 5.4 | **100** | 4.6 |
| | JM-sep($l=1,2; k=1$) | 5.5 | 6.0 | 5.0 | . | 6.6 | 5.4 | **100** | . |
| | JM-sep($l=1,2; k=2$) | 5.5 | 6.0 | . | 4.4 | 6.6 | 5.4 | . | 5.2 |
| | JM-sep($l=1; k=1$) | 5.0 | . | 5.0 | . | 6.1 | . | **100** | . |
| | JM-sep($l=1; k=2$) | 5.0 | . | . | 5.1 | 6.1 | . | . | 4.6 |
| | JM-sep($l=2; k=2$) | . | 6.7 | . | 4.9 | . | 5.2 | . | 4.7 |
| SNP3<sup>1</sup><br>(MAF=40%) | | $\beta_{g,1}=0$ | $\beta_{g,2}=0$ | $\gamma_{g,1}=0$ | $\gamma_{g,2}=0$ | $\beta_{g,1}=0$ | $\beta_{g,2}=0$ | $\gamma_{g,1}=0$ | $\gamma_{g,2}=0$ |
|  | JM-cmp | 4.0 | 6.7 | 4.3 | 4.5 | 4.5 | 4.6 | 4.7 | 3.8 |
|  | JM-mis | 4.0 | 6.7 | 4.6 | 3.9 | 4.5 | 4.6 | 89.9 | 58.4 |
| | JM-sep($l=1,2; k=1$) | 4.0 | 6.7 | 4.7 | . | 4.5 | 4.6 | 89.7 | . |
| | JM-sep($l=1,2; k=2$) | 4.0 | 6.7 | . | 4.4 | 4.5 | 4.6 | . | 57.7 |
| | JM-sep($l=1; k=1$) | 4.1 | . | 4.6 | . | 4.8 | . | 89.7 | . |
| | JM-sep($l=1; k=2$) | 4.1 | . | . | 4.6 | 4.8 | . | . | 33.0 |
| | JM-sep($l=2; k=2$) | . | 6.5 | . | 4.8 | . | 4.4 | . | 56.4 |
| SNP4<br>(MAF=30%) | | $\beta_{g,1}=0$ | $\beta_{g,2}=0$ | $\gamma_{g,1}=0$ | $\gamma_{g,2}=0$ | $\beta_{g,1}=0$ | $\beta_{g,2}=0$ | $\gamma_{g,1}=0$ | $\gamma_{g,2}=0.7$ |
|  | JM-cmp | 4.8 | 6.3 | 3.9 | 4.9 | 5.4 | 4.8 | 6.0 | **100** |
|  | JM-mis | 4.8 | 6.3 | 4.9 | 4.7 | 5.4 | 4.8 | 5.4 | **100** |
| | JM-sep($l=1,2; k=1$) | 4.8 | 6.3 | 4.9 | . | 5.4 | 4.8 | 5.1 | . |
| | JM-sep($l=1,2; k=2$) | 4.8 | 6.3 | . | 5.1 | 5.4 | 4.8 | . | **99.9** |
| | JM-sep($l=1; k=1$) | 5.0 | . | 4.6 | . | 6.1 | . | 5.1 | . |
| | JM-sep($l=1; k=2$) | 5.0 | . | . | 5.0 | 6.1 | . | . | **93.9** |
| | JM-sep($l=2; k=2$) | . | 5.6 | . | 4.8 | . | 4.8 | . | **99.9** |
| SNP5<br>(MAF=20%) | | $\beta_{g,1}=0$ | $\beta_{g,2}=0$ | $\gamma_{g,1}=0$ | $\gamma_{g,2}=0$ | $\beta_{g,1}=0$ | $\beta_{g,2}=7$ | $\gamma_{g,1}=0$ | $\gamma_{g,2}=0.4$ |
|  | JM-cmp | 5.0 | 5.5 | 4.7 | 5.6 | 2.8 | **100** | 5.5 | **66.4** |
|  | JM-mis | 5.0 | 5.5 | 4.6 | 4.5 | 2.8 | **100** | 4.6 | **64.5** |
| | JM-sep($l=1,2; k=1$) | 5.0 | 5.5 | 5.2 | . | 2.8 | **100** | 4.8 | . |
| | JM-sep($l=1,2; k=2$) | 5.0 | 5.5 | . | 5.0 | 2.8 | **100** | . | **63.2** |
| | JM-sep($l=1; k=1$) | 5.1 | . | 5.3 | . | 2.8 | . | 4.6 | . |
| | JM-sep($l=1; k=2$) | 5.1 | . | . | 4.9 | 2.8 | . | . | **100** |
| | JM-sep($l=2; k=2$) | . | 5.6 | . | 5.4 | . | **100** | . | **64.9** |

<sup>1</sup>Except for JM-cmp, none of the analyses account for indirect genetic pathways via the longitudinal risk factor $U$ . This translates into elevated T1E in the direct genetic effects $\gamma_{g,k}$ for both time-to-T1DC traits.

Under the alternative multi-SNP *causal genetic* scenario (Table 2, right-hand side), type I errors for tests of each null *β*_*g*,*l*_ tend to be close to the nominal level of 5% for most analysis models (with exceptions for SNP2 and SNP5), while tests of the two SNPs with effects on intermediate *measured* longitudinal QT risk factors (SNP1 (*β*_g,1_) and SNP5 (*β*_g,2_)) reach 100% power for all the analysis models (Table 2, right-hand side). Tests of null *γ*_*g*,*k*_ by JM-cmp, JM-mis and JM-sep(*l*=1,2; *k*, with *k*=1 or 2) show overall good type I error control for all SNPs with direct and/or indirect effects via measured longitudinal QTs (*i.e.* all SNPs but SNP3). However, for SNP1 with indirect effects on both T1DC traits via HbA1c, tests of null *γ*_g,2_ exhibit liberal type I errors in JM-sep(*l*=2; *k*=2), which may be explained by bias in SNP1 effect on DN (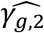, see File S2, section 6) towards the indirect SNP1 effect via HbA1c QT risk factor, which is ignored from JM-sep(*l*=2; *k*=2). For SNP2 which has direct effect on DR (*γ*_*g*,1_), tests of non-null *γ*_*g*,1_ exhibit equivalent or higher power under JM-cmp and JM-mis (closely followed by JM-sep(*l*=1,2; *k*)). However, the power to detect the direct SNP4 effect on DN (*γ*_*g*,2_) appears reduced in JM-sep(*l*=1; *k*=2); which can be explained by the bias in 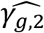 towards the null, due to ignoring SBP QT risk factor in JM-sep(*l*=1; *k*=2). For SNP5 with direct and indirect effects on DN via SBP, *γ*_*g*,2_ appears detected by a larger number of replicates under JM-sep(*l*=1; *k*=2) and CM-obs (Table 2, right-hand side); which may be explained by bias in SNP5 effect (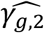, see File S2, section 6) towards the overall SNP5 effect (combining direct and indirect SNP5 effects via SBP), given that SBP is ignored from JM-sep(*l*=1; *k*=2) and random SBP variation in CM-obs. Finally, for SNP3 that has indirect effects on both T1DC traits via the *unmeasured* longitudinal risk factor *U*, only JM-cmp accounts for indirect SNP3 pathways via *U*. This translates into elevated type I errors for tests of each null *γ*_*g*,*k*_ likely explained by the biased SNP3 effects on both T1DC traits (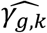, see File S2, section 6) in JM-mis, all JM-sep, and CM-obs, towards the indirect SNP3 effect via *U*. Overall, the relative ranking of empirical powers among the analysis models persists for tests of each *γ*_*g*,*k*_ across *P*\* varying up to 10^-5^ (File S2, section 8), with steeper power reduction for detection of SNP4 effect on DN (*γ*_*g*,2_) in JM-sep(*l*=1; *k*=2), and markedly misleading detection of SNP5 effect on DN (*γ*_*g*,2_) in JM-sep(*l*=1; *k*=2) and CM-obs.

As noted above, the improvement of the type I errors control and powers in tests of each *γ*_*g*,*k*_ under the alternative simulation scenario by JM-cmp (closely followed by JM-mis and JM-sep(*l*=1,2; *k=*1 or 2)) compared to JM-sep(*l*=1 or 2; *k=*1 or 2) for one longitudinal and one time-to-event trait and CM-obs can be explained by more efficient estimation accuracy in 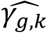, but also in the QT effect estimates on the TTE 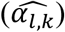 as illustrated by: bias reduction, coverage probabilities closer to the nominal level (95%), and empirical 95% confidence intervals of the parameter estimates narrower around the true parameter (File S2, section 6). Moreover, we find little evidence for correlation between 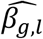 and 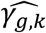. The average bootstrap correlation 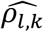 across replicates is low for each QT/TTE trait pair (*l*; *k*) under the *alternative genetic* (Tables 3-7) and *global null* scenarios (File S2, section 9) for all joint model analyses 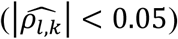. However, we see larger 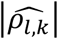 values in CM-obs, particularly for the SBP/DN trait pair; this may be explained by larger random variation in SBP which is ignored by CM-obs.

**Table 3.**
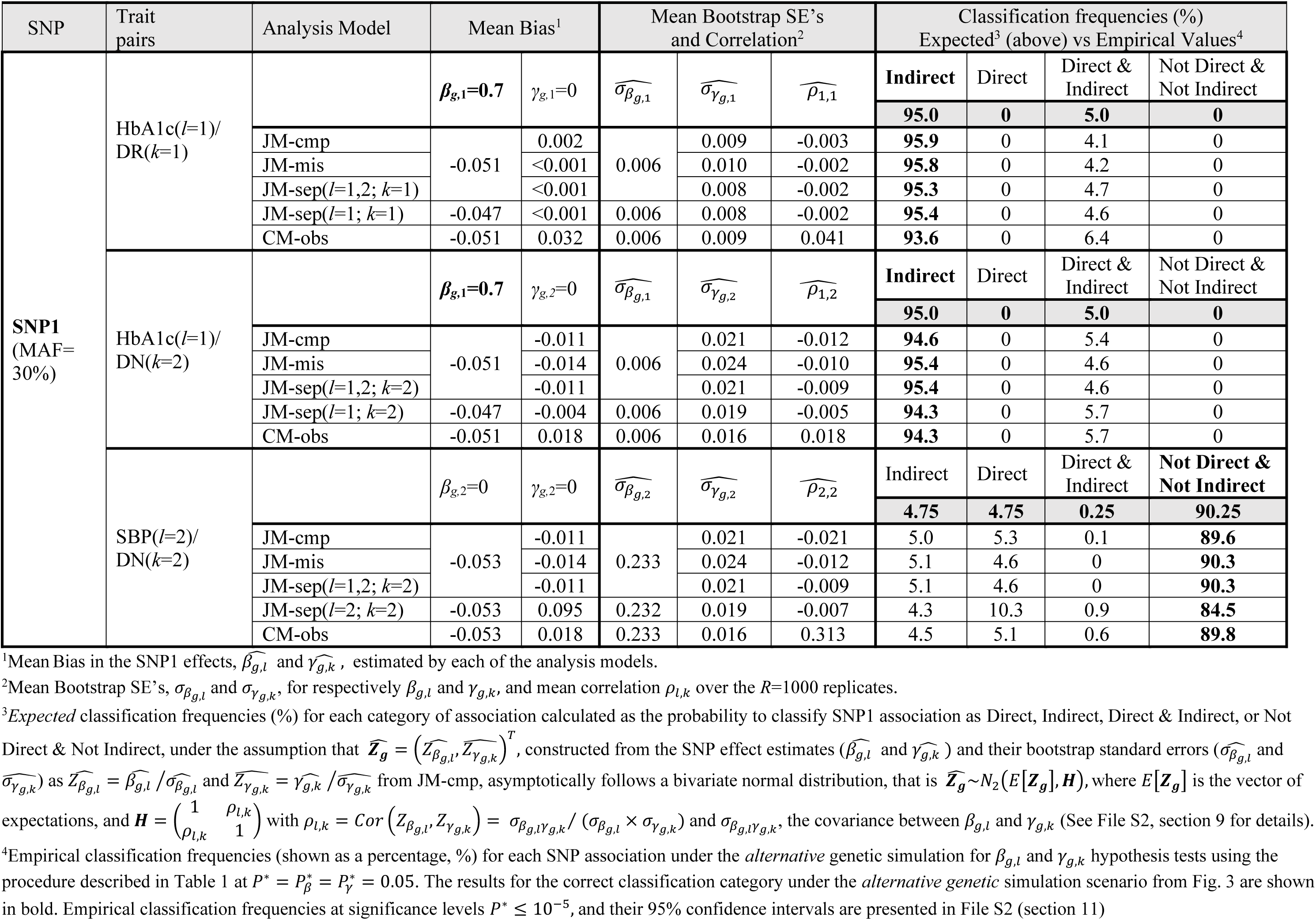
Classification frequencies for SNP1 association with each of the QT/time-to-event trait pairs based on the complete joint model and compared models at significance threshold *P*\*=0.05, using *R*=1000 replicates of *N*=667 DCCT subjects, with SNPs simulated under the *alternative* genetic scenario from Fig. 3

| SNP | Trait pairs | Analysis Model | Mean Bias<sup>1</sup> |  | Mean Bootstrap SE's and Correlation<sup>2</sup> |  |  | Classification frequencies (%) Expected<sup>3</sup> (above) vs Empirical Values<sup>4</sup> |  |  |  |
| --- | --- | --- | --- | --- | --- | --- | --- | --- | --- | --- | --- |
| SNP1<br>(MAF=30%) | HbA1c($l=1$)/<br>DR($k=1$) | | $\beta_{g,1}=0.7$ | $\gamma_{g,1}=0$ | $\widehat{\sigma_{\beta_{g,1}}}$ | $\widehat{\sigma_{\gamma_{g,1}}}$ | $\widehat{\rho_{1,1}}$ | **Indirect** | Direct | Direct & Indirect | Not Direct & Not Indirect |
|  |  |  |  |  |  |  |  | **95.0** | **0** | **5.0** | **0** |
|  |  | JM-cmp | -0.051 | 0.002 | 0.006 | 0.009 | -0.003 | **95.9** | 0 | 4.1 | 0 |
|  |  | JM-mis |  | <0.001 |  | 0.010 | -0.002 | **95.8** | 0 | 4.2 | 0 |
| | | JM-sep($l=1,2; k=1$) | | <0.001 | | 0.008 | -0.002 | **95.3** | 0 | 4.7 | 0 |
| | | JM-sep($l=1; k=1$) | -0.047 | <0.001 | 0.006 | 0.008 | -0.002 | **95.4** | 0 | 4.6 | 0 |
|  |  | CM-obs | -0.051 | 0.032 | 0.006 | 0.009 | 0.041 | **93.6** | 0 | 6.4 | 0 |
| | HbA1c($l=1$)/<br>DN($k=2$) | | $\beta_{g,1}=0.7$ | $\gamma_{g,2}=0$ | $\widehat{\sigma_{\beta_{g,1}}}$ | $\widehat{\sigma_{\gamma_{g,2}}}$ | $\widehat{\rho_{1,2}}$ | **Indirect** | Direct | Direct & Indirect | Not Direct & Not Indirect |
|  |  |  |  |  |  |  |  | **95.0** | **0** | **5.0** | **0** |
|  |  | JM-cmp | -0.051 | -0.011 | 0.006 | 0.021 | -0.012 | **94.6** | 0 | 5.4 | 0 |
|  |  | JM-mis |  | -0.014 |  | 0.024 | -0.010 | **95.4** | 0 | 4.6 | 0 |
| | | JM-sep($l=1,2; k=2$) | | -0.011 | | 0.021 | -0.009 | **95.4** | 0 | 4.6 | 0 |
| | | JM-sep($l=1; k=2$) | -0.047 | -0.004 | 0.006 | 0.019 | -0.005 | **94.3** | 0 | 5.7 | 0 |
|  |  | CM-obs | -0.051 | 0.018 | 0.006 | 0.016 | 0.018 | **94.3** | 0 | 5.7 | 0 |
| | SBP($l=2$)/<br>DN($k=2$) | | $\beta_{g,2}=0$ | $\gamma_{g,2}=0$ | $\widehat{\sigma_{\beta_{g,2}}}$ | $\widehat{\sigma_{\gamma_{g,2}}}$ | $\widehat{\rho_{2,2}}$ | Indirect | Direct | Direct & Indirect | **Not Direct & Not Indirect** |
|  |  |  |  |  |  |  |  | **4.75** | **4.75** | **0.25** | **90.25** |
|  |  | JM-cmp | -0.053 | -0.011 | 0.233 | 0.021 | -0.021 | 5.0 | 5.3 | 0.1 | **89.6** |
|  |  | JM-mis |  | -0.014 |  | 0.024 | -0.012 | 5.1 | 4.6 | 0 | **90.3** |
| | | JM-sep($l=1,2; k=2$) | | -0.011 | | 0.021 | -0.009 | 5.1 | 4.6 | 0 | **90.3** |
| | | JM-sep($l=2; k=2$) | -0.053 | 0.095 | 0.232 | 0.019 | -0.007 | 4.3 | 10.3 | 0.9 | **84.5** |
|  |  | CM-obs | -0.053 | 0.018 | 0.233 | 0.016 | 0.313 | 4.5 | 5.1 | 0.6 | **89.8** |

<sup>1</sup>Mean Bias in the SNP1 effects, $\widehat{\beta_{g,l}}$ and $\widehat{\gamma_{g,k}}$ , estimated by each of the analysis models.
<sup>2</sup>Mean Bootstrap SE's, $\widehat{\sigma_{\beta_{g,l}}}$ and $\widehat{\sigma_{\gamma_{g,k}}}$ , for respectively $\beta_{g,l}$ and $\gamma_{g,k}$ , and mean correlation $\rho_{l,k}$ over the $R=1000$ replicates.
<sup>3</sup>*Expected* classification frequencies (%) for each category of association calculated as the probability to classify SNP1 association as Direct, Indirect, Direct & Indirect, or Not Direct & Not Indirect, under the assumption that $\widehat{\mathbf{Z}}_g = (\widehat{Z_{\beta_{g,l}}}, \widehat{Z_{\gamma_{g,k}}})^T$ , constructed from the SNP effect estimates ( $\widehat{\beta_{g,l}}$ and $\widehat{\gamma_{g,k}}$ ) and their bootstrap standard errors ( $\widehat{\sigma_{\beta_{g,l}}}$ and $\widehat{\sigma_{\gamma_{g,k}}}$ ) as $\widehat{Z_{\beta_{g,l}}} = \widehat{\beta_{g,l}} / \widehat{\sigma_{\beta_{g,l}}}$ and $\widehat{Z_{\gamma_{g,k}}} = \widehat{\gamma_{g,k}} / \widehat{\sigma_{\gamma_{g,k}}}$ from JM-cmp, asymptotically follows a bivariate normal distribution, that is $\widehat{\mathbf{Z}}_g \sim N_2(E[\mathbf{Z}_g], \mathbf{H})$ , where $E[\mathbf{Z}_g]$ is the vector of expectations, and $\mathbf{H} = \begin{pmatrix} 1 & \rho_{l,k} \\ \rho_{l,k} & 1 \end{pmatrix}$ with $\rho_{l,k} = Cor(Z_{\beta_{g,l}}, Z_{\gamma_{g,k}}) = \sigma_{\beta_{g,l}\gamma_{g,k}} / (\sigma_{\beta_{g,l}} \times \sigma_{\gamma_{g,k}})$ and $\sigma_{\beta_{g,l}\gamma_{g,k}}$ , the covariance between $\beta_{g,l}$ and $\gamma_{g,k}$ (See File S2, section 9 for details).
<sup>4</sup>Empirical classification frequencies (shown as a percentage, %) for each SNP association under the *alternative* genetic simulation for $\beta_{g,l}$ and $\gamma_{g,k}$ hypothesis tests using the procedure described in Table 1 at $P^* = P_{\beta}^* = P_{\gamma}^* = 0.05$ . The results for the correct classification category under the *alternative genetic* simulation scenario from Fig. 3 are shown in bold. Empirical classification frequencies at significance levels $P^* \leq 10^{-5}$ , and their 95% confidence intervals are presented in File S2 (section 11)

### Classification of direct and/or indirect SNP associations with time-to-event traits

Under the *global null* scenario, the empirical classification frequencies for direct and/or indirect SNP association with each T1DC trait at significance level *P*;^*^ =0.05 agree with the expected classification frequencies for all the categories of SNP associations and for all the models (File S2, section 9); this observation confirms the accuracy of the proposed hypothesis procedure under the *global null* genetic scenario. We also find the expected classification frequencies to be insensitive to larger bootstrap correlation values 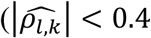, see File S2, section 9). Overall, empirical and expected classification frequencies are close to the nominal rate of 5% for the categories of direct or indirect association, but conservative for the category of direct and indirect association (see File S2, section 10); which requires rejection of single-parameter hypothesis for the two tests of SNP effects *β*_*g*,*l*_ and *γ*_*g*,*k*_ (Table1).

Under the *alternative* simulation scenario, when SNPs have direct and/or indirect effects via measured longitudinal QTs, we find that the proposed multivariate joint models, JM-cmp, JM-mis (and JM-sep(*l*=1,2; *k* with *k*=1 or 2)) lead to improved classification accuracy (empirical classification frequencies closer to the expected ones for each of the four categories of association) at specified significance level *P*\* = 0.05, and correctly classify the simulated SNP associations for each trait pair in more than 88% of replicates for SNP1, SNP2 and SNP4 (Tables 3 to 5), and in more than 61% of replicates for SNP5 (Table 6). In contrast, JM-sep(*l*=1 or 2; *k*=1 or 2) and CM-obs models exhibit larger differences between empirical and expected classification frequencies, which suggests lower ability of these models to correctly distinguish between direct and/or indirect association; which can lead to misleading inference. This is especially the case for the SNPs with bias in 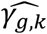 and low type I error control or power for test of 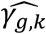 in JM-sep(*l*=1 or 2; *k*=1 or 2) and CM-obs models, as illustrated for:

- SNP1 that has indirect association with both T1DC traits via HbA1c; it exhibits lower-than-expected empirical classification frequencies for the correct association category with HbA1c/DR (indirect association) and with SBP/DN (not direct and not indirect association), see Table 3;
- SNP4 that has a direct association with DN, with lower than expected empirical classification frequency for the correct direct association category with HbA1c/DN in JM- sep(*l*=1; *k*=2) as shown in Table 5;
- SNP5 with direct and indirect effects on DN via SBP, which exhibit larger than expected empirical classification frequencies for the correct association category with HbA1c/DN (direct association), and with SBP/DN (direct and indirect association), see Table 6.

**Table 4.**
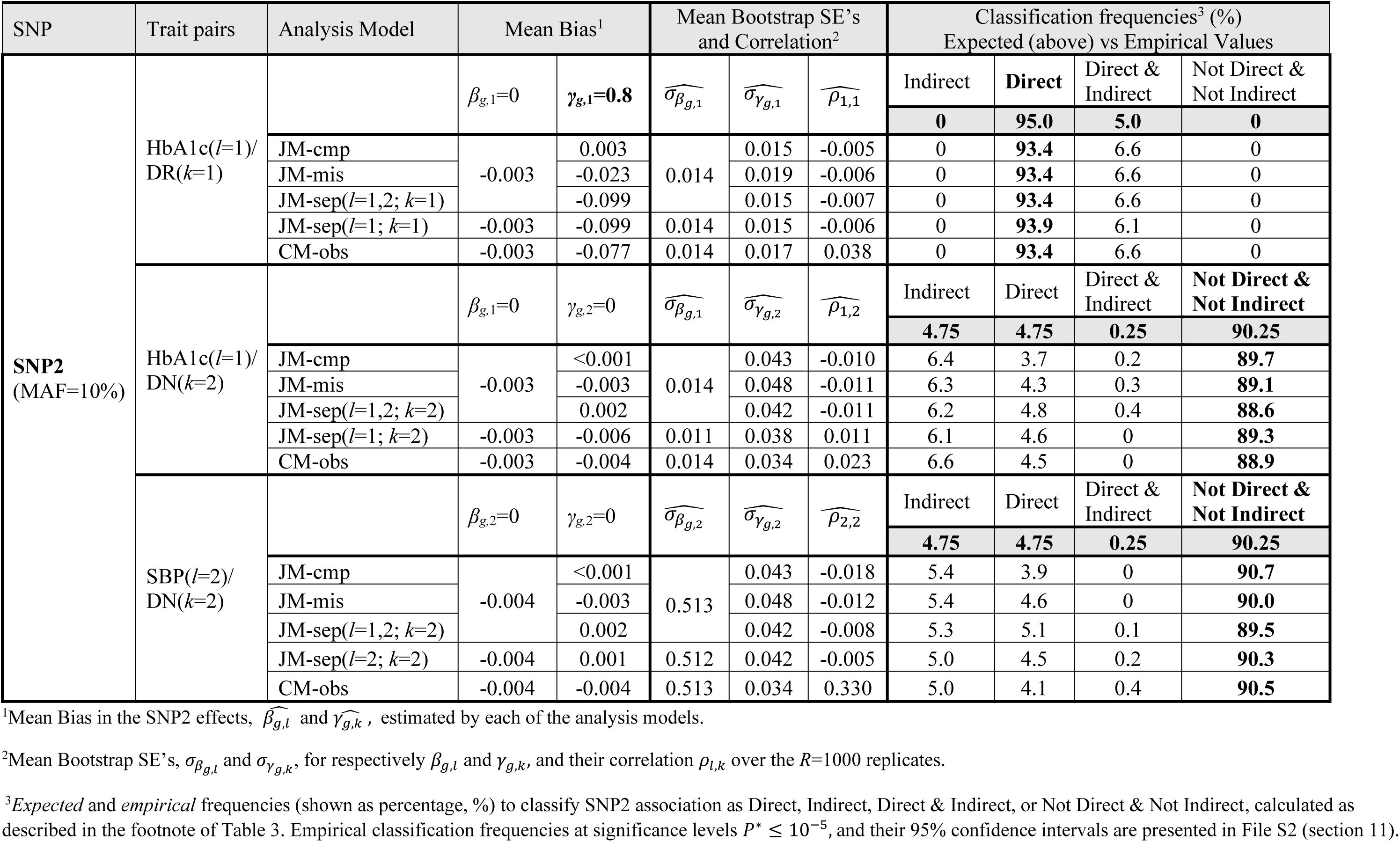
Classification frequencies for SNP2 association with each QT/time-to-event trait pairs based on the complete joint model and compared models at significance threshold P*=0.05, using R=1000 replicates of N=667 DCCT subjects, with SNPs simulated under the alternative genetic scenario from Fig. 3

| SNP | Trait pairs | Analysis Model | Mean Bias<sup>1</sup> |  | Mean Bootstrap SE's and Correlation<sup>2</sup> |  |  | Classification frequencies<sup>3</sup> (%)<br>Expected (above) vs Empirical Values |  |  |  |
| --- | --- | --- | --- | --- | --- | --- | --- | --- | --- | --- | --- |
| | | | $\beta_{g,1}=0$ | $\gamma_{g,1}=0.8$ | $\widehat{\sigma_{\beta_{g,1}}}$ | $\widehat{\sigma_{\gamma_{g,1}}}$ | $\widehat{\rho_{1,1}}$ | Indirect | Direct | Direct & Indirect | Not Direct & Not Indirect |
| SNP2<br>(MAF=10%) | HbA1c($l=1$)/<br>DR($k=1$) | | | | | | | **0** | **95.0** | **5.0** | **0** |
|  |  | JM-cmp | -0.003 | 0.003 | 0.014 | 0.015 | -0.005 | 0 | **93.4** | 6.6 | 0 |
|  |  | JM-mis |  | -0.023 |  | 0.019 | -0.006 | 0 | **93.4** | 6.6 | 0 |
| | | JM-sep($l=1,2; k=1$) | | -0.099 | | 0.015 | -0.007 | 0 | **93.4** | 6.6 | 0 |
| | | JM-sep($l=1; k=1$) | -0.003 | -0.099 | 0.014 | 0.015 | -0.006 | 0 | **93.9** | 6.1 | 0 |
|  |  | CM-obs | -0.003 | -0.077 | 0.014 | 0.017 | 0.038 | 0 | **93.4** | 6.6 | 0 |
| | HbA1c($l=1$)/<br>DN($k=2$) | | $\beta_{g,1}=0$ | $\gamma_{g,2}=0$ | $\widehat{\sigma_{\beta_{g,1}}}$ | $\widehat{\sigma_{\gamma_{g,2}}}$ | $\widehat{\rho_{1,2}}$ | Indirect | Direct | Direct & Indirect | Not Direct & Not Indirect |
|  |  |  |  |  |  |  |  | **4.75** | **4.75** | **0.25** | **90.25** |
|  |  | JM-cmp | -0.003 | <0.001 | 0.014 | 0.043 | -0.010 | 6.4 | 3.7 | 0.2 | **89.7** |
|  |  | JM-mis |  | -0.003 |  | 0.048 | -0.011 | 6.3 | 4.3 | 0.3 | **89.1** |
| | | JM-sep($l=1,2; k=2$) | | 0.002 | | 0.042 | -0.011 | 6.2 | 4.8 | 0.4 | **88.6** |
| | | JM-sep($l=1; k=2$) | -0.003 | -0.006 | 0.011 | 0.038 | 0.011 | 6.1 | 4.6 | 0 | **89.3** |
|  |  | CM-obs | -0.003 | -0.004 | 0.014 | 0.034 | 0.023 | 6.6 | 4.5 | 0 | **88.9** |
| | SBP($l=2$)/<br>DN($k=2$) | | $\beta_{g,2}=0$ | $\gamma_{g,2}=0$ | $\widehat{\sigma_{\beta_{g,2}}}$ | $\widehat{\sigma_{\gamma_{g,2}}}$ | $\widehat{\rho_{2,2}}$ | Indirect | Direct | Direct & Indirect | Not Direct & Not Indirect |
|  |  |  |  |  |  |  |  | **4.75** | **4.75** | **0.25** | **90.25** |
|  |  | JM-cmp | -0.004 | <0.001 | 0.513 | 0.043 | -0.018 | 5.4 | 3.9 | 0 | **90.7** |
|  |  | JM-mis |  | -0.003 |  | 0.048 | -0.012 | 5.4 | 4.6 | 0 | **90.0** |
| | | JM-sep($l=1,2; k=2$) | | 0.002 | | 0.042 | -0.008 | 5.3 | 5.1 | 0.1 | **89.5** |
| | | JM-sep($l=2; k=2$) | -0.004 | 0.001 | 0.512 | 0.042 | -0.005 | 5.0 | 4.5 | 0.2 | **90.3** |
|  |  | CM-obs | -0.004 | -0.004 | 0.513 | 0.034 | 0.330 | 5.0 | 4.1 | 0.4 | **90.5** |

<sup>1</sup>Mean Bias in the SNP2 effects, $\widehat{\beta_{g,l}}$ and $\widehat{\gamma_{g,k}}$ , estimated by each of the analysis models.
<sup>2</sup>Mean Bootstrap SE's, $\sigma_{\beta_{g,l}}$ and $\sigma_{\gamma_{g,k}}$ , for respectively $\beta_{g,l}$ and $\gamma_{g,k}$ , and their correlation $\rho_{l,k}$ over the $R=1000$ replicates.
<sup>3</sup>*Expected* and *empirical* frequencies (shown as percentage, %) to classify SNP2 association as Direct, Indirect, Direct & Indirect, or Not Direct & Not Indirect, calculated as described in the footnote of Table 3. Empirical classification frequencies at significance levels $P^* \leq 10^{-5}$ , and their 95% confidence intervals are presented in File S2 (section 11).

**Table 5.** Classification frequencies for SNP4 association with each pair of QT/time-to-event traits based on the complete joint model and compared models at significance threshold *P*\*=0.05, using *R*=1000 replicates of *N*=667 DCCT subjects, with SNPs simulated under the *alternative* genetic scenario from Fig. 3

| SNP | Trait pairs | Analysis Model | Mean Bias<sup>1</sup> |  | Mean Bootstrap SE's and Correlation<sup>2</sup> |  |  | Classification frequencies<sup>3</sup> (%)<br>Expected (above) vs Empirical Values |  |  |  |
| --- | --- | --- | --- | --- | --- | --- | --- | --- | --- | --- | --- |
| SNP4<br>(MAF=30%) | HbA1c($l=1$)/<br>DR($k=1$) | | $\beta_{g,1}=0$ | $\gamma_{g,1}=0$ | $\widehat{\sigma_{\beta_{g,1}}}$ | $\widehat{\sigma_{\gamma_{g,1}}}$ | $\widehat{\rho_{1,1}}$ | Indirect | Direct | Direct & Indirect | Not Direct & Not Indirect |
|  |  |  |  |  |  |  |  | **4.75** | **4.75** | **0.25** | **90.25** |
|  |  | JM-cmp | 0.001 | -0.001 | 0.006 | 0.007 | <0.001 | 4.9 | 5.5 | 0.5 | **89.1** |
|  |  | JM-mis |  | 0.001 |  | 0.009 | -0.003 | 5.0 | 5.0 | 0.4 | **89.6** |
| | | JM-sep($l=1,2; k=1$) | | 0.001 | | 0.007 | -0.003 | 5.1 | 4.8 | 0.3 | **89.8** |
| | | JM-sep($l=1; k=1$) | 0.001 | 0.001 | 0.006 | 0.007 | -0.002 | 5.7 | 4.7 | 0.4 | **89.2** |
|  |  | CM-obs | 0.001 | 0.001 | 0.006 | 0.008 | 0.038 | 5.0 | 4.2 | 0.4 | **90.4** |
| | HbA1c($l=1$)/<br>DN($k=2$) | | $\beta_{g,1}=0$ | $\gamma_{g,2}=0.7$ | $\widehat{\sigma_{\beta_{g,1}}}$ | $\widehat{\sigma_{\gamma_{g,2}}}$ | $\widehat{\rho_{1,2}}$ | Indirect | **Direct** | Direct & Indirect | Not Direct & Not Indirect |
|  |  |  |  |  |  |  |  | **0** | **94.98** | **5.0** | **0.02** |
|  |  | JM-cmp | 0.001 | -0.014 | 0.006 | 0.017 | -0.010 | 0 | **94.6** | 5.4 | 0 |
|  |  | JM-mis |  | -0.028 |  | 0.019 | -0.010 | 0 | **94.6** | 5.4 | 0 |
| | | JM-sep($l=1,2; k=2$) | | -0.076 | | 0.017 | -0.009 | 0 | **94.5** | 5.4 | 0.1 |
| | | JM-sep($l=1; k=2$) | 0.001 | -0.281 | 0.006 | 0.015 | 0.007 | 0.3 | **88.1** | 5.8 | 5.8 |
|  |  | CM-obs | 0.001 | -0.216 | 0.006 | 0.013 | 0.025 | 0 | **93.5** | 5.4 | 1.1 |
| | SBP($l=2$)/<br>DN($k=2$) | | $\beta_{g,2}=0$ | $\gamma_{g,2}=0.7$ | $\widehat{\sigma_{\beta_{g,2}}}$ | $\widehat{\sigma_{\gamma_{g,2}}}$ | $\widehat{\rho_{2,2}}$ | Indirect | **Direct** | Direct & Indirect | Not Direct & Not Indirect |
|  |  |  |  |  |  |  |  | **0** | **94.98** | **5.0** | **0.02** |
|  |  | JM-cmp | 0.012 | -0.014 | 0.215 | 0.017 | -0.026 | 0 | **95.2** | 4.8 | 0 |
|  |  | JM-mis |  | -0.028 |  | 0.019 | -0.021 | 0 | **95.2** | 4.8 | 0 |
| | | JM-sep($l=1,2; k=2$) | | -0.076 | | 0.017 | -0.020 | 0 | **95.1** | 4.8 | 0.1 |
| | | JM-sep($l=2; k=2$) | 0.012 | -0.088 | 0.215 | 0.016 | -0.019 | 0 | **95.1** | 4.8 | 0.1 |
|  |  | CM-obs | 0.012 | -0.216 | 0.215 | 0.013 | 0.332 | 0.1 | **94.2** | 4.7 | 1.0 |

<sup>1</sup>Mean Bias in the SNP4 effects, $\widehat{\beta_{g,l}}$ and $\widehat{\gamma_{g,k}}$ , estimated by each of the analysis models.
<sup>2</sup>Mean Bootstrap SE's, $\sigma_{\beta_{g,l}}$ and $\sigma_{\gamma_{g,k}}$ , for respectively $\beta_{g,l}$ and $\gamma_{g,k}$ , and their correlation $\rho_{l,k}$ over the $R=1000$ replicates.
<sup>3</sup>*Expected* and *empirical* frequencies (shown as percentage, %) to classify SNP4 association as Direct, Indirect, Direct & Indirect, or Not Direct & Not Indirect, calculated as described in the footnote of Table 3. Empirical classification frequencies at significance levels $P^* \leq 10^{-5}$ , and their 95% confidence intervals are presented in File S2 (section 11).

**Table 6.** Classification frequencies for SNP5 association with each pair of QT/time-to-event traits based on the complete joint model and compared models at significance threshold P*=0.05, using R=1000 replicates of N=667 DCCT subjects, with SNPs simulated under the alternative genetic scenario from Fig. 3

| SNP | Trait pairs | Analysis Model | Mean Bias<sup>1</sup> |  | Mean Bootstrap SE's and Correlation<sup>2</sup> |  |  | Classification frequencies<sup>3</sup> (%)<br>Expected (above) vs Empirical Values |  |  |  |
| --- | --- | --- | --- | --- | --- | --- | --- | --- | --- | --- | --- |
| | | | $\beta_{g,1}=0$ | $\gamma_{g,1}=0$ | $\widehat{\sigma_{\beta_{g,1}}}$ | $\widehat{\sigma_{\gamma_{g,1}}}$ | $\widehat{\rho_{1,1}}$ | Indirect | Direct | Direct & Indirect | Not Direct & Not Indirect |
| SNP5<br>(MAF=20%) | HbA1c($l=1$)/<br>DR($k=1$) | | | | | | | **4.75** | **4.75** | **0.25** | **90.25** |
|  |  | JM-cmp | -0.021 | <0.001 | 0.010 | 0.012 | 0.001 | 2.4 | 5.1 | 0.4 | **92.1** |
|  |  | JM-mis |  | 0.017 |  | 0.015 | <0.001 | 2.4 | 4.2 | 0.4 | **93.0** |
| | | JM-sep($l=1,2; k=1$) | | 0.013 | | 0.012 | <0.001 | 2.4 | 4.4 | 0.4 | **92.8** |
| | | JM-sep($l=1; k=1$) | -0.019 | 0.013 | 0.010 | 0.012 | -0.003 | 2.6 | 4.4 | 0.2 | **92.8** |
|  |  | CM-obs | -0.021 | 0.019 | 0.010 | 0.013 | 0.043 | 2.4 | 4.4 | 0.4 | **92.8** |
| | HbA1c($l=1$)/<br>DN($k=2$) | | $\beta_{g,1}=0$ | $\gamma_{g,2}=0.4$ | $\widehat{\sigma_{\beta_{g,1}}}$ | $\widehat{\sigma_{\gamma_{g,2}}}$ | $\widehat{\rho_{1,2}}$ | Indirect | **Direct** | Direct & Indirect | Not Direct & Not Indirect |
|  |  |  |  |  |  |  |  | **1.44** | **67.67** | **3.56** | **27.33** |
|  |  | JM-cmp | -0.021 | -0.020 | 0.010 | 0.025 | -0.016 | 1.1 | **64.7** | 1.7 | 32.5 |
|  |  | JM-mis |  | -0.009 |  | 0.029 | -0.013 | 0.8 | **62.5** | 2.0 | 34.7 |
| | | JM-sep($l=1,2; k=2$) | | -0.036 | | 0.026 | -0.014 | 1.2 | **61.6** | 1.6 | 35.6 |
| | | JM-sep($l=1; k=2$) | -0.019 | **0.765** | 0.010 | 0.021 | -0.008 | 0 | **97.2** | 2.8 | 0 |
|  |  | CM-obs | -0.021 | **0.505** | 0.010 | 0.022 | 0.017 | 0 | **97.2** | 2.8 | 0 |
| | SBP($l=2$)/<br>DN($k=2$) | | $\beta_{g,2}=7$ | $\gamma_{g,2}=0.4$ | $\widehat{\sigma_{\beta_{g,2}}}$ | $\widehat{\sigma_{\gamma_{g,2}}}$ | $\widehat{\rho_{2,2}}$ | Indirect | Direct | **Direct & Indirect** | Not Direct & Not Indirect |
|  |  |  |  |  |  |  |  | **28.77** | **0** | **71.23** | **0** |
|  |  | JM-cmp | 0.046 | -0.020 | 0.385 | 0.025 | -0.050 | 33.6 | 0 | **66.4** | 0 |
|  |  | JM-mis |  | -0.009 |  | 0.029 | -0.047 | 35.5 | 0 | **64.5** | 0 |
| | | JM-sep($l=1,2; k=2$) | | -0.036 | | 0.026 | -0.047 | 36.8 | 0 | **63.2** | 0 |
| | | JM-sep($l=2; k=2$) | 0.039 | -0.029 | 0.384 | 0.025 | -0.041 | 35.1 | 0 | **64.9** | 0 |
|  |  | CM-obs | 0.046 | **0.505** | 0.385 | 0.022 | 0.280 | 0 | 0 | **100** | 0 |

<sup>1</sup>Mean Bias in the SNP5 effects, $\widehat{\beta_{g,l}}$ and $\widehat{\gamma_{g,k}}$ , estimated by each of the analysis models.
<sup>2</sup>Mean Bootstrap SE's, $\sigma_{\beta_{g,l}}$ and $\sigma_{\gamma_{g,k}}$ , for respectively $\beta_{g,l}$ and $\gamma_{g,k}$ , and their correlation $\rho_{l,k}$ over the $R=1000$ replicates.
<sup>3</sup>*Expected* and *empirical* frequencies (shown as percentage, %) to classify SNP5 association as Direct, Indirect, Direct & Indirect, or Not Direct & Not Indirect, calculated as described in the footnote of Table 3. Empirical classification frequencies at significance levels $P^* \leq 10^{-5}$ , and their 95% confidence intervals are presented in File S2 (section 11).

On the other hand, for SNP3 which has indirect effects on both T1DC traits via the unmeasured longitudinal QT risk factor (*U*); except JM-cmp which exhibits accurate (and high) classification frequencies, all the other compared models (which ignore *U*), exhibit poor classification accuracy (Table 7) and tend to mistakenly classify SNP3 as a direct association with each T1DC trait in ∼30-86% of the replicates.

**Table 7.**
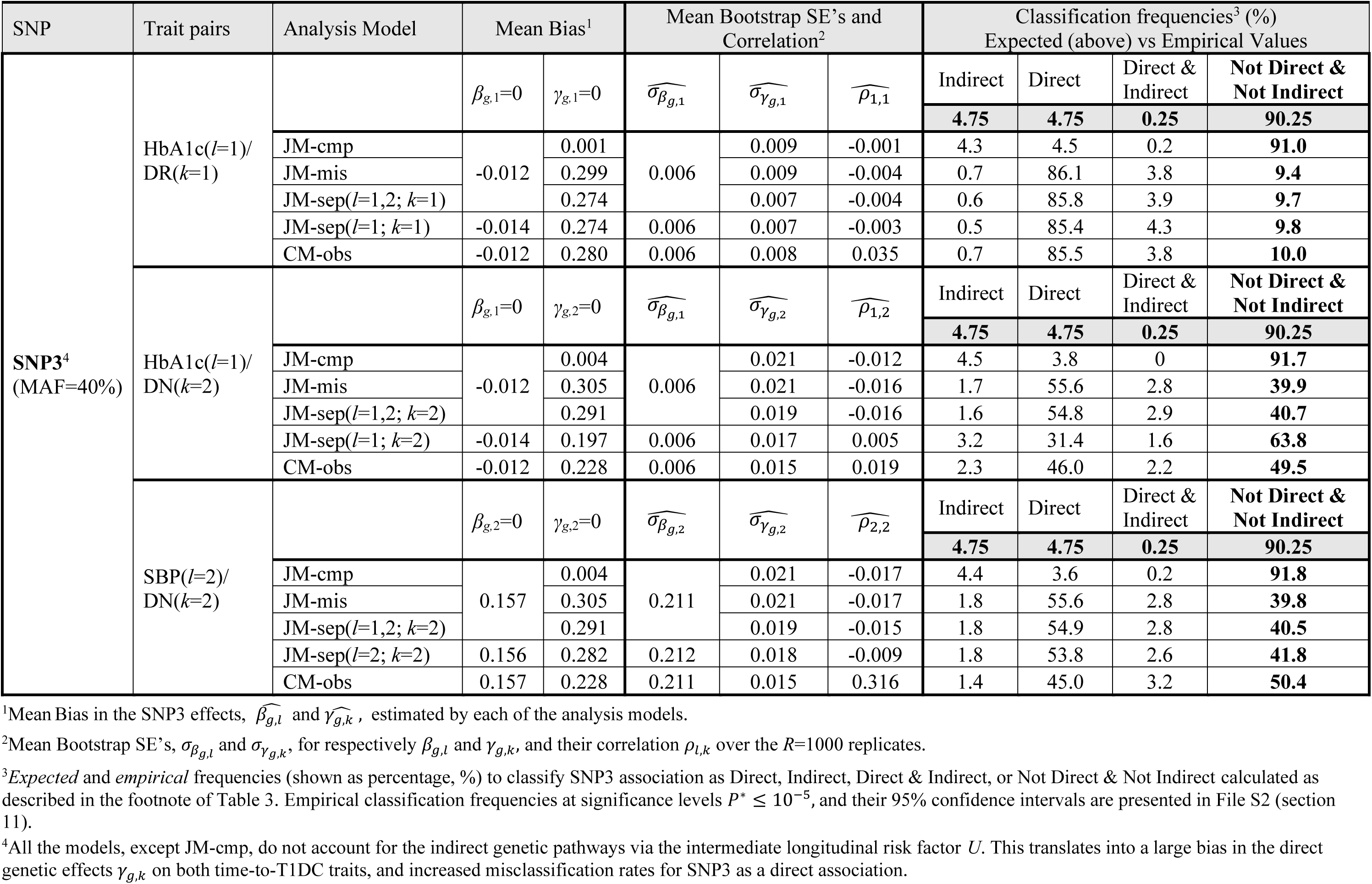
Classification frequencies for SNP3 association with each pair of QT/time-to-event traits based on the complete joint model and compared models at significance threshold *P*\*=0.05, using *R*=1000 replicates of *N*=667 DCCT subjects, with SNPs simulated under the *alternative* genetic scenario from Fig. 3

| SNP | Trait pairs | Analysis Model | Mean Bias<sup>1</sup> |  | Mean Bootstrap SE's and Correlation<sup>2</sup> |  |  | Classification frequencies<sup>3</sup> (%)<br>Expected (above) vs Empirical Values |  |  |  |
| --- | --- | --- | --- | --- | --- | --- | --- | --- | --- | --- | --- |
| | | | $\beta_{g,1}=0$ | $\gamma_{g,1}=0$ | $\widehat{\sigma}_{\beta_{g,1}}$ | $\widehat{\sigma}_{\gamma_{g,1}}$ | $\widehat{\rho}_{1,1}$ | Indirect | Direct | Direct & Indirect | Not Direct & Not Indirect |
| SNP3<sup>4</sup><br>(MAF=40%) | HbA1c($l=1$)/<br>DR($k=1$) | | | | | | | **4.75** | **4.75** | **0.25** | **90.25** |
|  |  | JM-cmp | -0.012 | 0.001 | 0.006 | 0.009 | -0.001 | 4.3 | 4.5 | 0.2 | **91.0** |
|  |  | JM-mis |  | 0.299 |  | 0.009 | -0.004 | 0.7 | 86.1 | 3.8 | **9.4** |
| | | JM-sep($l=1,2; k=1$) | | 0.274 | | 0.007 | -0.004 | 0.6 | 85.8 | 3.9 | **9.7** |
| | | JM-sep($l=1; k=1$) | -0.014 | 0.274 | 0.006 | 0.007 | -0.003 | 0.5 | 85.4 | 4.3 | **9.8** |
|  |  | CM-obs | -0.012 | 0.280 | 0.006 | 0.008 | 0.035 | 0.7 | 85.5 | 3.8 | **10.0** |
| | HbA1c($l=1$)/<br>DN($k=2$) | | $\beta_{g,1}=0$ | $\gamma_{g,2}=0$ | $\widehat{\sigma}_{\beta_{g,1}}$ | $\widehat{\sigma}_{\gamma_{g,2}}$ | $\widehat{\rho}_{1,2}$ | Indirect | Direct | Direct & Indirect | Not Direct & Not Indirect |
|  |  |  |  |  |  |  |  | **4.75** | **4.75** | **0.25** | **90.25** |
|  |  | JM-cmp | -0.012 | 0.004 | 0.006 | 0.021 | -0.012 | 4.5 | 3.8 | 0 | **91.7** |
|  |  | JM-mis |  | 0.305 |  | 0.021 | -0.016 | 1.7 | 55.6 | 2.8 | **39.9** |
| | | JM-sep($l=1,2; k=2$) | | 0.291 | | 0.019 | -0.016 | 1.6 | 54.8 | 2.9 | **40.7** |
| | | JM-sep($l=1; k=2$) | -0.014 | 0.197 | 0.006 | 0.017 | 0.005 | 3.2 | 31.4 | 1.6 | **63.8** |
|  |  | CM-obs | -0.012 | 0.228 | 0.006 | 0.015 | 0.019 | 2.3 | 46.0 | 2.2 | **49.5** |
| | SBP($l=2$)/<br>DN($k=2$) | | $\beta_{g,2}=0$ | $\gamma_{g,2}=0$ | $\widehat{\sigma}_{\beta_{g,2}}$ | $\widehat{\sigma}_{\gamma_{g,2}}$ | $\widehat{\rho}_{2,2}$ | Indirect | Direct | Direct & Indirect | Not Direct & Not Indirect |
|  |  |  |  |  |  |  |  | **4.75** | **4.75** | **0.25** | **90.25** |
|  |  | JM-cmp | 0.157 | 0.004 | 0.211 | 0.021 | -0.017 | 4.4 | 3.6 | 0.2 | **91.8** |
|  |  | JM-mis |  | 0.305 |  | 0.021 | -0.017 | 1.8 | 55.6 | 2.8 | **39.8** |
| | | JM-sep($l=1,2; k=2$) | | 0.291 | | 0.019 | -0.015 | 1.8 | 54.9 | 2.8 | **40.5** |
| | | JM-sep($l=2; k=2$) | 0.156 | 0.282 | 0.212 | 0.018 | -0.009 | 1.8 | 53.8 | 2.6 | **41.8** |
|  |  | CM-obs | 0.157 | 0.228 | 0.211 | 0.015 | 0.316 | 1.4 | 45.0 | 3.2 | **50.4** |

<sup>1</sup>Mean Bias in the SNP3 effects, $\widehat{\beta}_{g,l}$ and $\widehat{\gamma}_{g,k}$ , estimated by each of the analysis models.
<sup>2</sup>Mean Bootstrap SE's, $\sigma_{\beta_{g,l}}$ and $\sigma_{\gamma_{g,k}}$ , for respectively $\beta_{g,l}$ and $\gamma_{g,k}$ , and their correlation $\rho_{l,k}$ over the $R=1000$ replicates.
<sup>3</sup>*Expected* and *empirical* frequencies (shown as percentage, %) to classify SNP3 association as Direct, Indirect, Direct & Indirect, or Not Direct & Not Indirect calculated as described in the footnote of Table 3. Empirical classification frequencies at significance levels $P^* \leq 10^{-5}$ , and their 95% confidence intervals are presented in File S2 (section 11).
<sup>4</sup>All the models, except JM-cmp, do not account for the indirect genetic pathways via the intermediate longitudinal risk factor $U$ . This translates into a large bias in the direct genetic effects $\gamma_{g,k}$ on both time-to-T1DC traits, and increased misclassification rates for SNP3 as a direct association.

As stringency of *P*;^*^ increases up to 10^-5^ for the same effect sizes, empirical classification frequencies decrease in the correct association category, while mis-classification frequencies increase in the other categories; for example SNP5 tends to be mistakenly classified as an indirect association with SBP/DN with JM-cmp (File S2, section 11).

In summary, our simulations show that by using smoothed trajectories and accounting for all QT associations, the extended joint model improves parameter estimation efficiency and classification accuracy of SNPs directly associated with each time-to-event trait or indirectly associated via a measured longitudinal QT in comparison to classification using JM-sep models for analysis of each time-to-event trait with one or multiple longitudinal traits or CM-obs. Reduced classification accuracy translate to increased risk of misclassifying a SNP as direct and/or indirect association. In addition, when a SNP has both direct and indirect effects on a time-to-event trait, the proposed classification procedure can be conservative because it requires the joint significance of the two SNP effects, *β*_*g*,*l*_ and *γ*_*g*,*k*_, where the power of each test depends on effect size, MAF and trait distribution. As a result, a SNP with both direct and indirect associations can be misclassified as either a direct or an indirect association, as we will also illustrate in the application results in DCCT. Finally, when a SNP has an indirect effect on both T1DC traits via an unmeasured QT, the testing procedure based on JM-mis, that captures some of the unmodeled dependency between time-to-event traits through the frailty term, does not prevent misclassification as a direct association, which could be due to the limitation of using a time-invariant frailty term. In addition, this observation also demonstrates the importance of the proposed joint model extension, which allows analysis on all the intermediate QT(s), as opposed to JM-sep, to avoid misclassification of direct and/or indirect SNP associations.

### Application in the DCCT Genetics Study data

We demonstrate feasibility of the proposed two-stage extended joint model method by analysis in the DCCT Genetics Study data. The application is based on DCCT individuals from the Conventional treatment group genotyped on HumanCoreExome Bead Array (Illumina, San Diego, CA, USA) with ungenotyped autosomal SNPs imputed using 1000 Genomes data phase 3 (The 1000 Genomes Project Consortium 2015), as detailed in File S1. We use time to mild retinopathy and time to persistent microalbuminuria, for DR and DN outcomes respectively, as previously defined in the motivating GWAS of HbA1c (Paterson et al. 2010); see File S3, section 1 for details. Out of 667 conventionally-treated DCCT individuals with genetic data, we analyze *N*=516 subjects, excluding those with mild to moderate non-proliferative retinopathy or DN at DCCT baseline. By the time of the DCCT close-out visit, 297 (57.6%) had a DR event, and 61 (11.8%) had a DN event, including 47 subjects (9.1%) that experienced both events. After SNP filtering and pruning on linkage disequilibrium (File S3, section 2), we analyze 307 SNPs reported as associated with HbA1c, SBP, or multiple definitions of DR and/or DN (Paterson et al. 2010; Grassi et al. 2011; Sandholm et al. 2012; Hosseini et al. 2015; Wheeler et al. 2017; Evangelou et al. 2018; Pollack et al. 2019), see File S4 for the full list of SNPs.

#### Joint model fitting

We fit the joint model for each SNP one at a time, including baseline covariates (age at diagnosis, T1D duration, cohort, sex, and year of DCCT study entry), longitudinal HbA1c and SBP, and T1DCs DR and DN (File S3, section 3). Given prior evidence for long term HbA1c effects on T1DC, we present results under the time-weighted cumulative for HbA1c association with time-to-T1DC traits, which exhibits stronger prior association with T1DC in the DCCT individuals analyzed here, but we obtained similar classification results under the two alternative specifications considered for HbA1c association (i.e. contemporaneous value and updated mean, see File S3, section 4). We apply diagnostic tools in the joint model analysis, including residual analysis in the longitudinal and time-to-event sub-models, finding little evidence for model misspecification (File S3, section 5). In the Cox PH frailty time-to-event sub-model, PH assumptions are well-satisfied when the baseline hazard is stratified on the cohort variable; overall conclusions are equivalent when cohort is included as a covariate. As shown in File S3 (section 5), the martingale residuals of the Cox PH frailty model are consistent with the assumption of a linear relationship between the QTs and each time-to-T1DC outcome.

In Fig. 5 (Panel A), we show that rs1358030 and rs10810632, which were discovered in a previous GWAS of HbA1c in DCCT (Paterson et al. 2010), are classified as indirect associations with both T1DC traits via their association with the HbA1c shared risk factor (*P*;_*βg*_≤ *P*;^*^ and *P*;_*γ*_ > *P*;^*^, *P*;^*^*=*1.7×10^-4^ after Bonferroni correction for the 289.02 effective SNPs tested (Li and Ji 2005)). Although the other candidate SNPs were selected from larger meta-analysis in T1D and/or independent studies in general populations (Paterson et al. 2010; Grassi et al. 2011; Sandholm et al. 2012; Hosseini et al. 2015; Wheeler et al. 2017; Evangelou et al. 2018; Pollack et al. 2019), the SNP-trait association tests did not reach Bonferroni-corrected significance thresholds in our analysis (File S3, section 4), potentially due to effect heterogeneity and low power to detect these variants or by some inherent heterogeneity factors in the study designs and/or phenotypic definitions. Thus, classification for these candidate SNPs is uninformative.

**Fig. 5.**
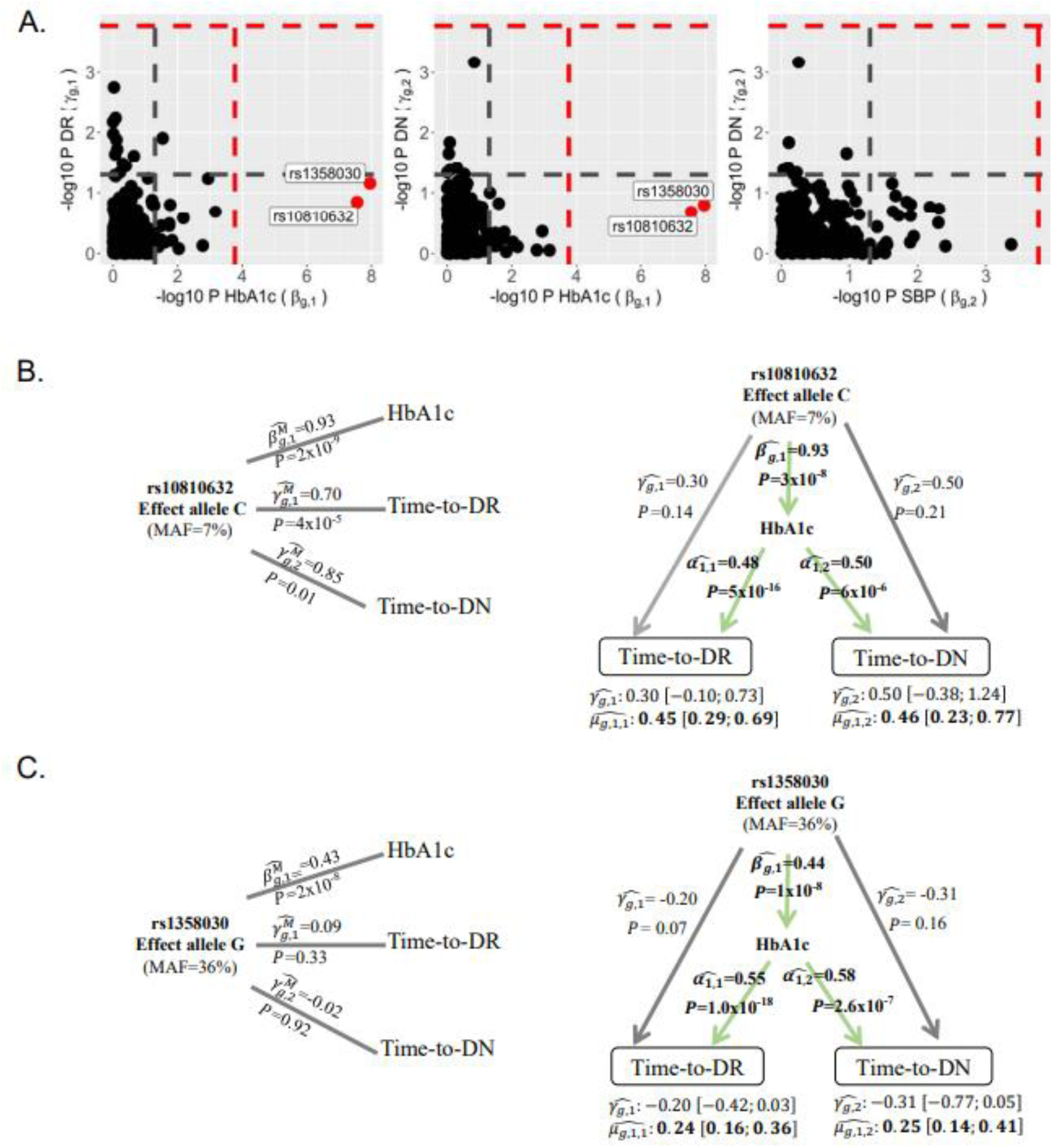
Classification of direct and/or indirect SNP associations in the DCCT Genetics Study data. (**A**) Scatter plots of the *P*-values (-log10) for tests of *β*_*g*,*l*_ (H0: *β*_*g*,*l*_ = 0 vs H1 *β*_*g*,*l*_ ≠ 0) on the *X* axis and *γ*_*g*,*k*_ (H0: *γ*_*g*,*k*_ = 0 vs H1: *γ*_*g*,*k*_ ≠ 0) on the *Y* axis for HbA1c/DR, HbA1c/DN and SBP/DN trait pairs. Significance levels *P*;^*^ =1.7×10^-4^ and *P*;^*^ = 0.05 are indicated by red and grey horizontal and vertical dashed lines. (**B**) and (**C**) represent association results for rs10810632 and rs1358030 detected as indirect associations at *P*;^*^ =1.7×10^-4^. Left panels present results from separate analysis of each trait (*ie.* longitudinal model for each QT and Cox PH time-to-event model without adjusting for the longitudinal traits) as used in naïve discovery GWAS; and right panels show results from the joint model with bootstrap 95% confidence intervals for the direct and indirect SNP effects. Results are presented using time-weighted cumulative HbA1c effects on T1DC traits.

#### Sample size and power considerations

The DCCT-based simulation study shows that multiple-trait joint model analysis can achieve high classification rates for direct and/or indirect SNP associations under SNP effect sizes in the specified range (SNP effects: on the QTs 0.7-7, on T1DC 0.4-0.8; and MAFs 10-40%) and classification rates under the global null hypothesis (null SNP effects on all traits) controlled to the nominal levels for direct or indirect SNP association but conservative for direct and indirect association. Compared to the simulation, the number of DN events is lower in the DCCT data application. We thus expect lower power to detect direct SNP association with DN; this implies reduced accuracy to classify a SNP as having either a direct, or a direct and indirect association.

In light of the simulation results, we acknowledge that a larger sample size would be required to maintain performance in DCCT for genetic associations with lower MAFs/effect sizes than the ones specified. In particular, lower effect sizes and MAFs can lead to larger variances of estimated effects and larger 95% confidence intervals, as observed for the direct effects of rs10810632 (MAF=7%) compared to rs1358030 (MAF=36%) for both time-to-T1DC traits (Fig. 5, Panels B and C). SNP effects with larger variances have lower power to detect association. Although we expect the effect sizes estimated by the joint model to be relatively insensitive to the study sample size, reduction in variances by increasing study sample size would be expected to improve classification accuracy for these two SNPs (particularly for rs10810632 that has a lower MAF).

To assess how genetic association and classification results for rs10810632 and rs1358030 may be affected by increasing sample size, we applied parametric resampling (see File S3, section 6 for details) to draw datasets with sample size up to five times the DCCT sample of *N*=516, and we then extrapolated the classification beyond *N*=2580. This analysis demonstrates decreasing variances of the SNP effects for both SNPs as sample size increases and narrowing of the 95% and 99% confidence intervals (File S3, section 6); panel A in Fig. 6 illustrates the corresponding shift in classification of rs10810632 association with DR as indirect via HbA1c towards classification as both indirect and direct. On the other hand, given that both SNPs were previously discovered in a GWAS of HbA1c in DCCT, SNP effect estimates for HbA1c association 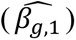 may be overestimated due to Winner’s curse bias (Kraft 2008; Sun et al. 2011), which would impact the classification results. We therefore repeated the sample size analyses specifying an adjusted SNP effect size for the HbA1c association equal to 50% of its effect estimate in our DCCT analysis. In this scenario the test of SNP association with HbA1c falls just below the *P*\* threshold in the resample size *N*=516, although power improves in larger sample sizes (Fig. 6). Here again classification tends to shift from indirect towards both direct and indirect SNP associations with both T1DC traits (see File S3, section 6 for complete results for both SNPs); however, a much larger sample size is needed to cross the *P\** threshold for test of direct association.

**Fig. 6.**
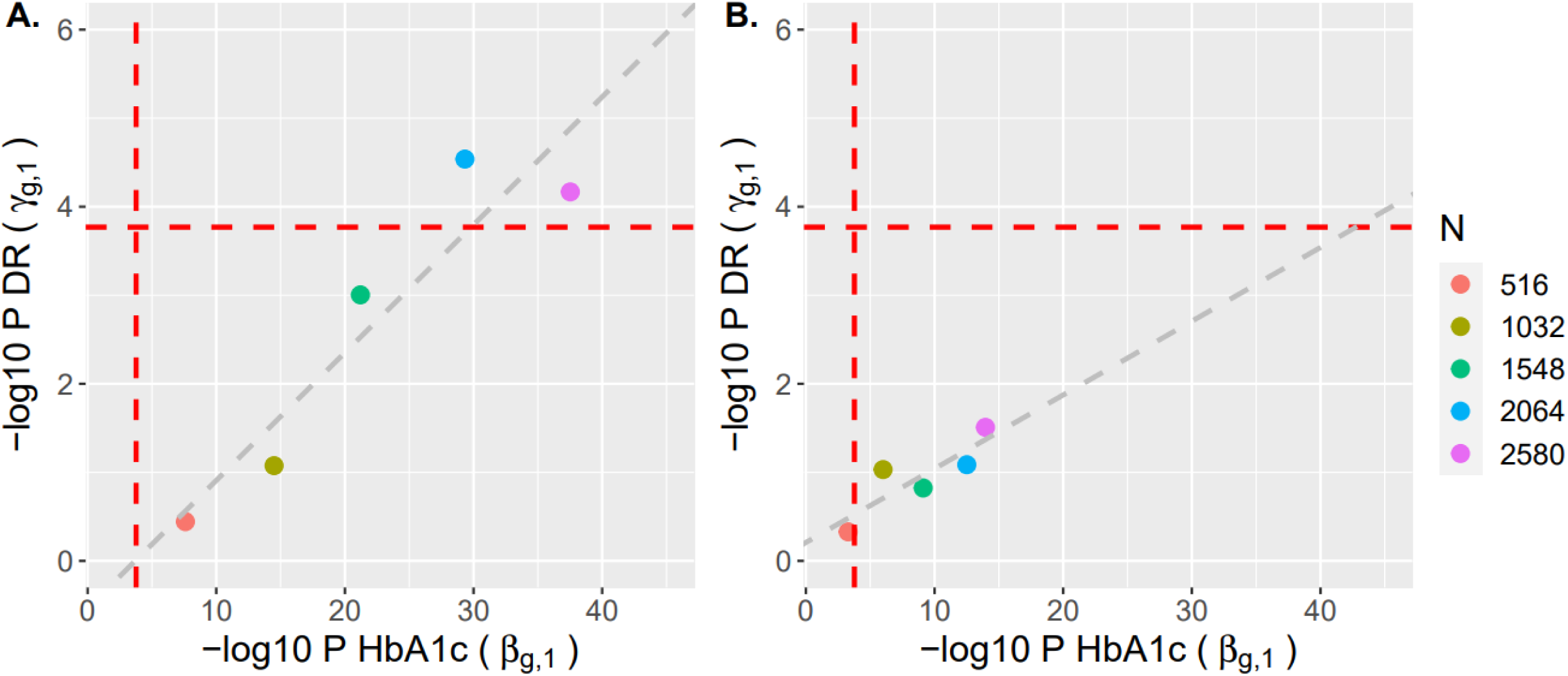
Change in classification results for rs10810632 with HbA1c/DR to increasing sample size (A) and to the Winner’s curse bias for the SNP effect on HbA1c (B) investigated using parametric resampling. We used parametric resampling (File S3, section 6) to draw datasets with sample size up to five times the DCCT sample size of *N*=516, and then extrapolated the classification results beyond *N*=2580. The *X* axes of (**A**) and (**B**), show the *P*-values (-log10) for the test of rs10810632 effect on HbA1c (H0: *β*_*g*,1_ = 0 vs H1 *β*_*g*,1_ ≠ 0)), while the *Y* axes show the *P*-values (-log10) for the test of rs10810632 effect on DR (H0: *γ*_*g*,1_ = 0 vs H1: *γ*_*g*,1_ ≠ 0) for the sample sizes investigated (shown by different colors). We fitted a regression line on each plot to project the trend of the classification beyond *N*=2580 individuals. These plots illustrate the corresponding shift in classification of rs10810632 association with DR as indirect via HbA1c towards classification as both indirect and direct association. Complete results with HbA1c/DN are shown in File S3 (section 6).

In the DCCT application, we take advantage of the largest available clinical study of T1D complications with long-term follow-up and high-density longitudinal QT measurements. This highlights the dearth of longitudinal studies with both a large number of individuals and long-term clinical follow-up, as well as the related challenges in joint model analysis. In prospective study designs with both longitudinal and time-to-event traits, an inherent imbalance can exist among traits in detection of SNP associations, in that power for QT(s) depends on the number of measurements while power for time-to-event traits depends on the number of events and duration of follow-up. Although there is currently a trade-off between epidemiological studies with large sample sizes but low density of longitudinal measurements, and clinical studies with more modest sample sizes but high measurement densities, we anticipate informative application of joint model analysis in large biobanks, for example (Scholtens et al. 2015; Bycroft et al. 2018; Dummer et al. 2018), now assembling longitudinal measures jointly with binary outcomes and genetic data.

## DISCUSSION

We present new, more informative methods for statistical genetic analysis under a joint model specification of multiple longitudinal risk factors and multiple time-to-event traits, designed to characterize the complex genetic architecture of related traits in longitudinal studies of disease progression. The proposed extended model is formulated to deal with dependencies within and between traits and can account for trait-specific and shared covariates, within-subject random variability in the longitudinal traits, as well as effects of unobserved baseline confounding factors between the time-to-event traits through a subject-specific frailty term. We also introduce a realistic data-based simulation algorithm to assess joint model performance that can also be used to estimate achievable power in clinical studies such as DCCT characterized by extensive longitudinal follow-up but limited sample size.

Evaluation by realistic simulation study of complex T1DC genetic architecture shows that accounting for trait dependencies and measurement errors in longitudinal QT risk factors using the proposed joint model extension improves classification accuracy of SNP as direct and/or indirect association compared to *(i)* separate joint model analysis of each time-to-event trait with one or multiple longitudinal traits, and *(ii)* Cox-PH frailty model analysis adjusted for multiple observed longitudinal trait values. This improvement in classification accuracy under the joint models of multiple longitudinal and multiple TTE traits results from improved Type I error control for single-parameter tests of each of the two SNP effects (*β*_*g*,*l*_ or *γ*_*g*,*k*_), and improved power to detect SNP association, that can be explained by reduced estimation bias in parameters. However, we also observe that estimation bias and misclassification can be severe in the presence of SNP association with a longitudinal risk factor that is unmeasured or absent from the joint model, and mis-classification may be non-negligible when the study has limited power to detect either of the two SNP effects (*β*_*g*,*l*_ or *γ*_*g*,*k*_), as in the DCCT study. We apply parametric resampling to evaluate how study sample size or Winner’s curse bias affects classification accuracy and anticipate that this approach may also help inform replication study design with sufficient power. Nevertheless, we conclude that application of joint model analysis in longitudinal studies of disease progression, such as in the DCCT Genetics Study, improves classification of direct and/or indirect SNP association and helps to elucidate the genetic architecture of complex traits.

Although the primary aim in this report is to develop statistical methods to distinguish among direct and/or indirect SNP associations with each time-to-event trait, the multi-trait extension of the joint model lends itself to development of multi-trait SNP association testing for SNP discovery. In File S5, we present a joint-parameter test for global SNP association with all the longitudinal and time-to-event traits, based on a generalized Wald statistic. In application to the simulated DCCT-based complex genetic architecture, we observe good type I error control under the global genetic null scenario, improved power for SNP discovery when a SNP has multiple trait effects, and power maintenance in other SNP association scenarios.

Although the extended joint model can be applied to studies where longitudinal QT measurements are missing at some of the visits, choice of the joint model estimation method depends on the missing data mechanism. In the presence of informative missing data mechanisms, we recommend sensitivity analysis using existing implementations of joint likelihood estimation that assume the time-to-dropout mechanism depends on missing longitudinal QT values through the posterior distribution of the random effects; this corresponds to an informative missing data mechanism (Rizopoulos 2012). To our knowledge these implementations exist only for simple joint model formulations with either one longitudinal and one time-to-event trait (Rizopoulos 2010) or with multiple longitudinal traits and one time-to-event outcome (Rizopoulos 2016; Hickey et al. 2018c). Alternatively multiple imputation methods have been implemented for two-stage estimation (Rubin 1987; Moreno-Betancur et al. 2018); these impute missing values for multiple longitudinal continuous traits using the conditional distribution of each QT trait given one time-to-event trait and other QTs. More generally for missing data, further development of methods to maximize the extended joint likelihood, for example using Bayesian methods, would alleviate numerical challenges with increasing model complexity in multivariate extensions of joint models (Lawrence Gould et al. 2015); but this would require the design of an efficient sampling algorithm to study the posterior distribution.

We acknowledge several features of the joint model approach that warrant examination in further work. *Firstly*, to reduce computational complexity and improve model flexibility, we use two-stage parameter estimation. In some circumstances, this approach can produce biased estimates in the longitudinal and/or time-to-event sub-models as well as underestimation of their standard errors (Tsiatis et al. 1995; Wulfsohn and Tsiatis 1997; Dafni and Tsiatis 1998; Ye et al. 2008; Albert and Shih 2010; Sweeting and Thompson 2011; Ye and Wu 2017; Huong et al. 2018; Mauff et al. 2020). Biased estimates can result from informatively missing data in the presence of non-random censoring of the longitudinal trait values due to the occurrence of an event or from informative dropout (Ye et al. 2008; Albert and Shih 2010; Mauff et al. 2020). The simulation results show minimal biases in the absence of informative censoring, even when an associated longitudinal QT variable is omitted from the joint model. In the DCCT application, characterized by administrative censoring and a high completion rate, these biases are of less concern because longitudinal trait values continued to be recorded on a pre-specified visit schedule regardless of the occurrence of any T1DC events; we estimated the trajectories using all the available measurements. Furthermore, we obtain robust non-parametric bootstrap estimates of the covariance matrix for the SNP effects, and simulation results under the null do not show deviation from expected distributions. *Secondly*, because the joint model integrates longitudinal and time-to-event sub-models, model misspecification can occur in multiple ways and lead to invalid inference (Arisido et al. 2019). We recommend a careful assessment of model assumptions and data-based simulation studies to evaluate the robustness of classification of direct and/or indirect associations to two-stage assumptions. In the DCCT application, we provide an illustration of diagnostic analyses which could serve as guidance in other applications. *Thirdly*, patient visits were scheduled with high frequency in DCCT, so we ignored the modest degree of interval censoring in the current implementation of the joint model; when there are longer gaps between visits, extended methods are needed to account for interval censoring in the time-to-event sub-model with additional simulation studies to assess impact on joint model estimates.

*Computationally*, joint model fitting can be very demanding, particularly for genetic association studies that test millions of variants. In the DCCT application, it took ∼1 minute to fit the joint model for each SNP and ∼ 18 more minutes to estimate the covariance matrix with 500 bootstraps run in parallel on 4 nodes (each node with 40 CPU and 202 GB RAM). While analysis at the genome-wide level, involving for example ∼9 million imputed autosomal SNPs in DCCT Study (Roshandel et al. 2018), is computationally unrealistic at present, a screening approach without bootstrap to select informative SNPs, followed by bootstrap refinement would reduce the computational burden to feasible levels. Recently, computationally efficient algorithms have been developed to improve feasibility of linear mixed model (Sikorska et al. 2018) and Cox PH model (Rizvi et al. 2019; Bi et al. 2020) analyses for genome-wide genetic association studies, but to date, they remain to be implemented for multivariate outcomes. *Lastly*, (Liu et al. 2018) discuss various formulations and interpretations of joint models in the context of mediation analysis, with shared-random-effects accounting for potential unmeasured baseline confounding factors between one longitudinal and one time-to-event traits. Using applications in datasets from two clinical trials, they illustrate interpretation of sensitivity analysis to unmeasured baseline confounders. Adaptation of the joint model we propose for multiple longitudinal and multiple time-to-event traits for mediation analysis requires extension of the mediation assumptions (Sobel 1982; Mackinnon et al. 1995) to the case of multiple mediators and multiple time-to-event traits. Specific evaluations of the proposed model under these assumptions are also warranted.

We expect application of joint model methods in large biobank datasets to be informative in characterization of the genetic architecture of complex traits. Some extensions of joint models have been proposed to account for additional challenges raised by large biobanks, for example informative visiting processes (Gasparini et al. 2020). By providing more efficient SNP effect estimates and increased precision in polygenic risk score development, results of such analysis have potential to contribute to the translation of human genetic findings into personalized medicine (Young et al. 2019), as well as to causal inference using mediation and mendelian randomization studies. Finally, the joint model framework has potential to enable dynamic prediction beneficial for dynamic risk assessment (Papageorgiou et al. 2019; Bull et al. 2020) and optimization of intervention strategies (Sweeting and Thompson 2011; Yuen et al. 2018).

## Data Availability

DCCT data are available to authorized users at https://repository.niddk.nih.gov/studies/edic/ and https://www.ncbi.nlm.nih.gov/projects/gap/cgi-bin/study.cgi?study_id=phs000086.v3.p1 (IRB #07-0208-E). Example R codes for DCCT-data-based simulation and analysis of the simulated data are provided on GitHub (https://github.com/brossardMyriam/Joint-model-for-multiple-trait-genetics). Supplementary files are available on Figshare at https://figshare.com/s/2b9f6b3da5e1f03e8086. File S1 includes the description of the DCCT dataset as well as the list of the participants of the DCCT/EDIC Research Group; File S2 includes supplemental information for the DCCT-based simulation study; File S3 includes supplemental information for the Analysis of the DCCT Genetics Study data; File S4 includes the list of SNPs analyzed in DCCT; File S5 includes some notes on a multi-trait SNP association test for SNP effects estimated under the proposed joint model framework.

https://www.ncbi.nlm.nih.gov/projects/gap/cgi-bin/study.cgi?study_id=phs000086.v3.p1.

https://github.com/brossardMyriam/Joint-model-for-multiple-trait-genetics

## Acknowledgements

This study uses data provided by the Diabetes Control and Complications Trial / Epidemiology of Diabetes Interventions and Complications (DCCT/EDIC) Research Group which is sponsored through research contracts from the National Institute of Diabetes, Endocrinology and Metabolic Diseases of the National Institute of Diabetes and Digestive Kidney Diseases (NIDDK) and the National Institutes of Health (NIH). The authors are grateful to the subjects in the DCCT/EDIC cohort for their long-term participation. A complete list of the individuals and institutions participating in the DCCT/EDIC Research Group can be found in File S1. This project was supported by: CIHR Operating/Project Grants (#MOP-84287, #PJT-159509, #PJT-159463), CANSSI Collaborative Research Team in “Statistical methods for the analysis of genetic data with survival outcomes”, CANSSI postdoctoral fellowship (MB), CIHR STAGE fellowships (MB and OEG, #GET-101831). Computations were performed on the Niagara supercomputer at the SciNet HPC Consortium. SciNet is funded by the Canada Foundation for Innovation under the auspices of Compute Canada; the Government of Ontario; Ontario Research Fund - Research Excellence; and the University of Toronto.

## Abbreviations

DAG: directed acyclic graph
DCCT: Diabetes control and complications trial
DR: diabetic retinopathy
DN: diabetic nephropathy
GWAS: genome-wide association study
HbA1c: Hemoglobin A1c
LD: linkage disequilibrium
MAF: minor allele frequency
PH: proportional hazards
QT(s): quantitative trait(s)
SBP: systolic blood pressure
SNP: single nucleotide polymorphism
T1DC: type 1 diabetes complications
TTE: time-to-event

## Appendix

### Joint likelihood function of the joint model of one longitudinal and one time-to-event trait (L=K=1)

Under the following assumptions (A1-A3),

(A1) ***b_i_***∼*N*_2_(0, ***D***), where ***b_i_*** = (*b*_*i*,0_, *b*_*i*,1_)*^T^* are subject-specific random effects
(A2) *ε*_*i*,*j*_∼*N*(0, *σ*^2^) and *ε*_*i*,*j*_ ⊥ *ε*_*i*,*s*_ between visit times *t*_*i*,*j*_ ≠ *t*_*i*,*s*_, with *j* ≠ *s*, for all 1 ≤ *j* ≤ *J* and 1 ≤ *s* ≤ *J*
(A3) ***b_i_*** ⊥ ***ε_i_***, where ***ε_i_*** = (*ε*_*i*,1_, …, *ε*_*i*,*j*_, …, *ε*_*i*,*J*_)^*T*^, ***ε_i_***∼*N*_*J*_(0, *σ*^2^***I***_***J***_)

Then conditional on the random effects ***b_i_*** and fixed effects ***Ω***, it is appropriate to assume: the repeated measurements in the longitudinal process are independent (Laird and Ware 1982), in other words that the serial correlation is taken into account; and the longitudinal and time-to-event trait models are independent (Ibrahim et al. 2001). Under these conditional independence assumptions, the joint likelihood function of the joint model parameters ***Ω*** given the observed data is:

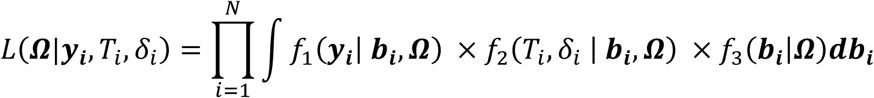

where:

- 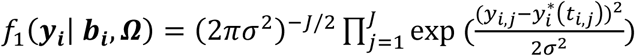
- *f*_2_(*T*_*i*_, *δ*_*i*_ | ***b_i_***, ***Ω***) = [*λ*_*i*_(*T*_*i*_| ***b_i_***, ***Ω***)]^*δi*^ *S*_*i*_(*T*_*i*_| ***b_i_***, ***Ω***), with
- 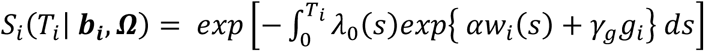 and 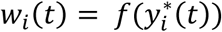 where 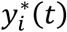, denotes the *l*^th^ QT trajectory at time *t* for 1 ≤ *l* ≤ *L* which depends on the fixed and random effects ***β*** and ***b_i_***. The survival function depends on the whole QT history of the true unobserved longitudinal process up to time *t*, noted as *Y*^*^(*t*)*=*{ *y*^*^(*s*), 0 ≤ *s* ≤ *t*};
- 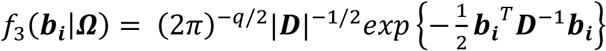, where *q* is the dimension of the ***D*** matrix.

The event indicator *δ*_*i*_ is used to distinguish the contribution of the individuals who experience the event during the observation period from the individuals who are still at risk up to that time point but do not experience the event. Individuals who experience the event (*δ*_*i*_ = 1) contribute to the cumulative hazard function and to the hazard function both evaluated at the *T*_*i*_; the individuals who do not experience the event (*δ*_*i*_ = 0) contribute to the hazard function only.

Joint model parameter estimation can be performed by maximization of the full joint likelihood function directly or by Bayesian computation. Direct maximization of the joint likelihood function can be performed using the Expectation-Maximization (EM) algorithm, treating the random effects as missing data (Wulfsohn and Tsiatis 1997). However, integrals with respect of time in the definition of the survival function, as well as the integral with respect to the ***b_i_*** do not have an analytical solution; therefore, numerical approaches, such as adaptive Gauss-Hermit quadrature, need to be used. Implementations of this model using a maximization of the above joint likelihood function have been proposed in different R packages (see (Furgal et al. 2019) for a review). However, as the dimension of the random effects increases, the integral over the random effects becomes computationally burdensome and Bayesian approaches can be employed instead, where the random effects are also considered model parameters obtained as a posterior sample and thus the integral over the random effects is no longer required.

### Joint model of multiple longitudinal and multiple time-to-event traits

#### Joint likelihood function

Extending the previous joint model assumptions (A1-A3) to *L*>1 and *K*>1, we have

(A1) ***b_i_***∼*N*_2_(0, ***D***), where ***b_i_*** = (***b***_*i*,**1**_, …, ***b***_*i*,***l***_, …, ***b***_*i*,***L***_)*^T^* are subject-specific random effects for all *L* QTs

(A2) ***ε***_*i*,***l***_∼*N*_*J*_(**0**, *σ*^2^***I***_***J***_) for all *l*^th^ QT with 1 ≤ *l* ≤ *L*

(A3) ***b***_*i*,***l***_ ⊥ ***ε***_*i*,***l***_ for all *l*^th^ QT with 1 ≤ *l* ≤ *L*, and

(A4) *u*_*i*_ ⊥ ***b***_*i*,***l***_, where *u*_*i*_ is a shared subject-specific frailty for *k* time-to-event traits.

Then, conditional on the random effects ***b_i_***, the frailty *u*_*i*_, and fixed effects ***Ω***, we further assume: ***b_i_*** accounts for association among the *L* longitudinal traits (Shah et al. 1997) and association between the longitudinal and time-to-event outcomes (Ibrahim et al. 2001); and the frailty term accounts for residual dependence among the time-to-event traits (Hougaard 1995). Under these conditional independence assumptions, the joint likelihood function of the joint model parameters ***Ω*** given the observed data is:

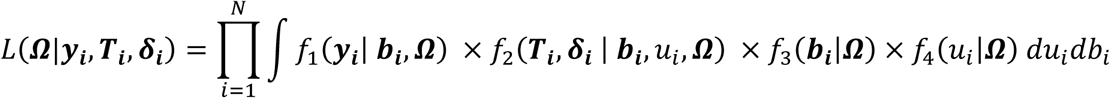

where:

- (***T***_*i*_, ***δ***_*i*_) = ((*T*_*i*,1_, *δ*_*i*,1_)*,…,* (*T*_*i*,*k*_, *δ*_*i*,*k*_),…,(*T*_*i*,*K*_, *δ*_*i*,*K*_))^*T*^ is defined as the vector of *K* stacked time-to-event outcomes for individual *i*,
- 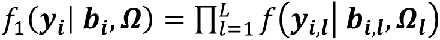 with 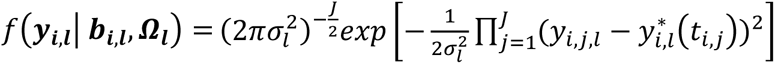
- 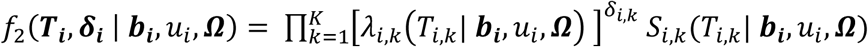, with 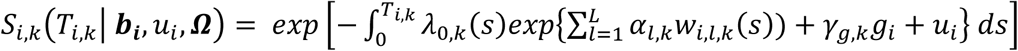, and 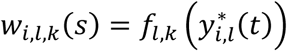, where 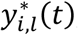, denotes the *l*^th^ QT trajectory at time *t* for 1 ≤ *l* ≤ *L* which depends on the fixed and random effects ***β***_***l***_ and ***b***_*i*,***l***_.
- 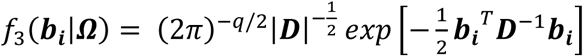, where *q* is the dimension of the ***D*** matrix
- 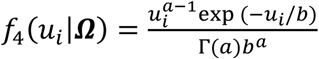, i.e. we assume *u*_*i*_∼Gamma(*a*, *b*) with *u*_*i*_ > 0, *a* corresponds to the shape parameter and *b* to the scale parameter, and *a*, *b*>0. Γ(*a*) is the gamma function evaluated at *a*.

We are not aware of any existing implementations of full likelihood maximization of the extended model in the literature. Calculation of the full likelihood requires multivariate integration with respect to the random effects distribution, which can lead to demanding computation. When the random effect vector ***b_i_***, has a small dimension, say less than 3, the integral can be evaluated via Gaussian quadrature which approximates the integral by a weighted sum of the target function evaluated at pre-specified sample points. However, when the dimension is larger, it is demanding to calculate the integrals with satisfactory approximation accuracy. Although a full likelihood specification enables rigorous study of asymptotic properties, its large sample approximation may not be accurate when sample size is small. In comparison, the Bayesian paradigm does not require asymptotic approximations, but the design of an efficient sampling algorithm to study the posterior distribution is challenging. Because of these limitations, we implement a two-stage approach to estimation of fixed effect parameters in the extended multi-trait model that is reasonable for the GWAS application of interest; in particular the longitudinal measurements in DCCT are taken according to a pre-specified schedule and are not terminated by the observation of diabetes complications, loss to follow-up and mortality are minimal, censoring is administrative, and each individual has a dense and nearly complete set of measurements.

### Likelihood functions under the two-stage approximation

Let ***ΩLong*** and ***ΩSurv*** be the vectors containing all fixed parameters from the longitudinal and time-to-event sub-models respectively.

***Stage 1***: Multivariate mixed model

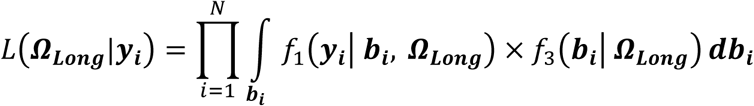

Where:

- 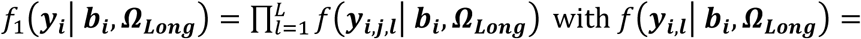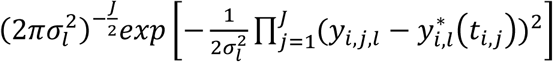
- 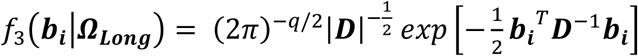 where *q* is the dimension of the ***D***

The fixed-effect and random-effect parameters are estimated jointly for all longitudinal traits using all available repeated measurements, without using the time-to-event information. Then fitted trajectory values are obtained by plugging the parameter estimates into:

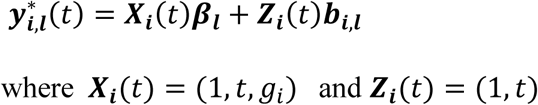

***Stage 2***: Multivariate Cox PH model adjusted for fitted trajectory values for the vector of *L* longitudinal outcomes

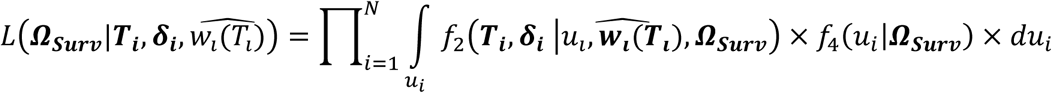

With:

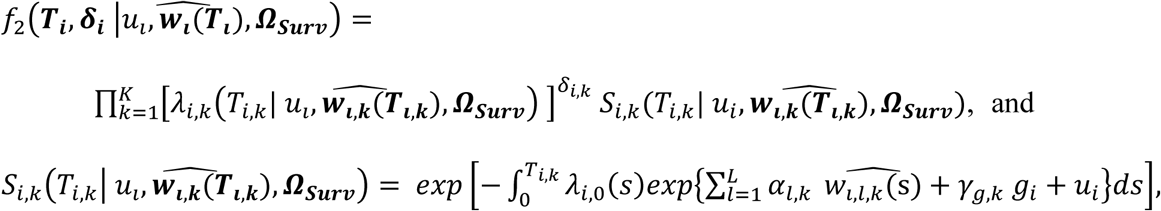

where 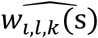 is obtained by plugging fitted trajectory values into 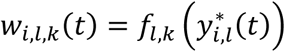.

Unlike the *joint likelihood function*, where the shared random effects ***b_i_*** account for the dependencies between the longitudinal and the time-to-event traits, the two-stage approach accounts for the dependencies between the longitudinal and time-to-event traits via the fitted values of the longitudinal trajectories. This approximation can produce biased estimates and/or underestimated standard errors for longitudinal and survival model parameters, when there is non-random censoring of the longitudinal trait values due to the occurrence of an event or from informative dropout (Ye et al. 2008; Albert and Shih 2010), and because of propagation errors of Stage 1 parameter estimates into Stage 2 (Wulfsohn and Tsiatis 1997). Under longitudinal model mis-specification and estimation bias, the conditional independence assumption can fail, undermining the accurate of trajectory estimates. Because the time-to-event processes are related to length of follow-up, informative missingness/dropouts can lead to differential follow-up between subjects with and without an event, and thus the random effects ***b_i_*** can depend on the event times (e.g. patients who have an event early are more likely to have positive random slopes). However, as we show in the simulation studies, in absence of model mis-specification and informative dropouts/missingness, this approach has low bias and is computationally feasible for genetic association studies.

